# Cross-disorder comparison of Brain Structures among 4,836 Individuals with Mental Disorders and Controls utilizing Danish population-based Clinical MRI Scans

**DOI:** 10.1101/2025.03.19.25324239

**Authors:** Stefano Cerri, Vardan Nersesjan, Kiril Vadimovic Klein, Enric Cristòbal Cóppulo, Sebastian Nørgaard Llambias, Mostafa Mehdipour Ghazi, Mads Nielsen, Michael Eriksen Benros

**Author notes:** shared last authorship (Mads Nielsen, Michael Eriksen Benros). **Correspondence:** - Stefano Cerri, PhD, Gentofte Hospitalsvej 19, st., 2900, Hellerup, Denmark, - Professor Michael E. Benros, MD, PhD, Gentofte Hospitalsvej 19, st., 2900, Hellerup, Denmark.

## Abstract

Large-scale mega-analyses of worldwide combined Magnetic Resonance Imaging (MRI) studies have demonstrated brain differences between individuals with mental disorders and controls. However, the potential of large-scale observational studies using population-based clinical MRI data remains unexplored. We analyzed clinical MRI data from 23,545 patients in the Eastern half of Denmark (Capital Region of Denmark and Region Zealand). 2,774 patients with mental disorders and 2,062 non-psychiatric controls fulfilled our inclusion and exclusion criteria. Patients with mental disorders exhibited smaller thalamic (d=−0.298) and amygdala volumes (d=−0.250), with larger ventricles (d=0.272), and thinner insula (d=−0.177), all p<0.0001. Analysis across all ROIs revealed a widespread pattern of thinner cortex (d=−0.180), especially in the temporal pole (d=−0.234) and superior frontal (d=−0.212) regions, and increased extracerebral cerebrospinal fluid (d=0.264). For volumetric measurements, findings were consistent across different inclusion and exclusion criteria but varied for cortical thickness measurements. Utilizing this currently largest population-based MRI cohort for mental disorders, we demonstrate that clinical MRI scans can detect brain structural differences among patients with mental disorders in real-world clinical settings, aiding in the stratification of patients without mental disorders. Cross-disorder analyses reveal shared neuroanatomical changes, including globally smaller brain volumes and thinner cortex. Integrating large-scale clinical MRI data with electronic health records holds promise for improved patient stratification and tracking of disease progression for future longitudinal cross-disorder studies, bridging real-world MRI data with clinical trajectories for further biological subgrouping.

## INTRODUCTION

Prior brain imaging studies aiming to identify brain patterns in mental disorders have often been constrained by small sample sizes with publication bias towards positive findings, highlighting the critical need for larger datasets to ensure reproducibility.^1,2^ Nevertheless, Magnetic Resonance Imaging (MRI) scans have proven effective in detecting subtle brain changes across various mental disorders in large-scale studies.^3^

To produce more reliable results, the pioneering consortium ENIGMA (Enhancing NeuroImaging Genetics through Meta-Analysis)^4^ pools neuroimaging data from multiple research centers worldwide to increase statistical power and generalize findings across different mental disorders. Despite their studies being the largest to date,^3, 5–10^ heterogeneity in imaging protocols, MRI processing, inclusion and exclusion criteria for patients and controls regarding diagnoses, treatments, and demographics across centers might introduce noise and hinder the generalizability of their findings. Additionally, the lack of comprehensive patient history information, in connection to the ENIGMA MRI data, constrains their ability to correlate brain findings with patient clinical data. Most importantly, as the ENIGMA consortia data is the largest dataset to date, validation of their findings in independent large-scale datasets is strongly needed.

Recent advances in automatic tools for extracting cortical and subcortical information from MRI scans have enabled the analysis of large-scale clinical MRI data,^11–13^ available in quantities far exceeding typical research study samples. Clinical MRI data, often accompanied by electronic health records (EHRs), allows for better patient stratification and more precise inclusion and exclusion criteria based on complete patient histories. Furthermore, as recent advances have shown shared clinical and genetic features across the spectrum of mental disorders,^14,15^ cross-disorder investigations are increasingly conducted within psychiatric research.^16–21^

Given these advancements, our study has two primary objectives: First, to assess whether prior findings based on high-quality MRI data can be replicated with routinely acquired clinical MRI scans. Second, to investigate the influence of inclusion and exclusion criteria based on medication and patient history on brain changes associated with mental disorders, both cross-disorder and within diagnostic subgroups. By addressing these objectives, we aim to highlight the potential of large-scale clinical MRI data for advancing the understanding of the neurobiological mechanisms underlying mental disorders and exploring the utility of current real-world MRI in contributing to diagnostic biomarkers.

## METHODS

We conducted a retrospective cohort study utilizing all clinical MRI scans from the Eastern half of Denmark (the Capital Region of Denmark and Region Zealand) acquired during the year 2019, totaling 313,927 MRI images obtained during 28,129 MRI visits on 23,545 unique patients. We first defined our cases with mental disorders and non-psychiatric control using complete EHRs available for all patients, including demographics, diagnoses, and medications. Secondly, anatomical brain regions of interest were defined based on prior large-scale studies and meta-analyses comparing mental disorder patients with controls, to investigate the reproducibility of previous findings. Lastly, we explored the robustness and dynamics of these differences by adding stricter exclusion criteria for cases and non-psychiatric controls. We obtained legal and ethical approvals for the proposed study from the Data Protection Agency and the Ethics Committee of the Capital Region of Denmark (R-22002033, and P-2020-101).

### Study Participants

Individuals ≥18 years of age with mental disorders were included as cases using the following ICD-10 codes: any Mental Disorders (F00-F99), and specifically Dementia (F00-F03 and G30), Substance Use Disorder (F10-19), Schizophrenia Spectrum Disorder (F20-29), Depression (F32-F33), and Anxiety Disorder (F40-F48). For patients with multiple of these diagnoses, a hierarchy was applied—Dementia, Schizophrenia Spectrum Disorder, Depression, Anxiety Disorder, and Substance Use Disorder—based on increasing ICD-10 code values. Substance Use Disorder was placed last to avoid overriding other psychiatric diagnoses and isolate the effect of substance abuse. Individuals had to be diagnosed with a mental disorder within three months of the MRI scan. This threshold was determined through sensitivity analysis (**Supplementary Figure S1**). Controls were ≥18 years of age and without mental or neurological disorders. These basic inclusion and exclusion criteria were termed “Population A”, similar to the minimum criteria to be included in ENIGMA consortium studies. “Population B” additionally included brain medications as exclusion criteria for non-psychiatric controls. Lastly, further exclusions were based on neurological medications and neurological comorbidities for cases (“Population C”). Details for each criteria are provided in **Figure 1**, and specific medication and diagnosis codes are listed in **Supplementary Methods**. Subgroup analyses included only diagnoses with ≥50 individuals meeting Population C criteria, to minimize false-positive findings and overestimation of effect sizes.^1,2^ Cross-disorder analyses included all mental disorders (F00–F99, G30) to investigate neuroanatomical patterns shared across the diagnostic spectrum.

**Figure 1:**
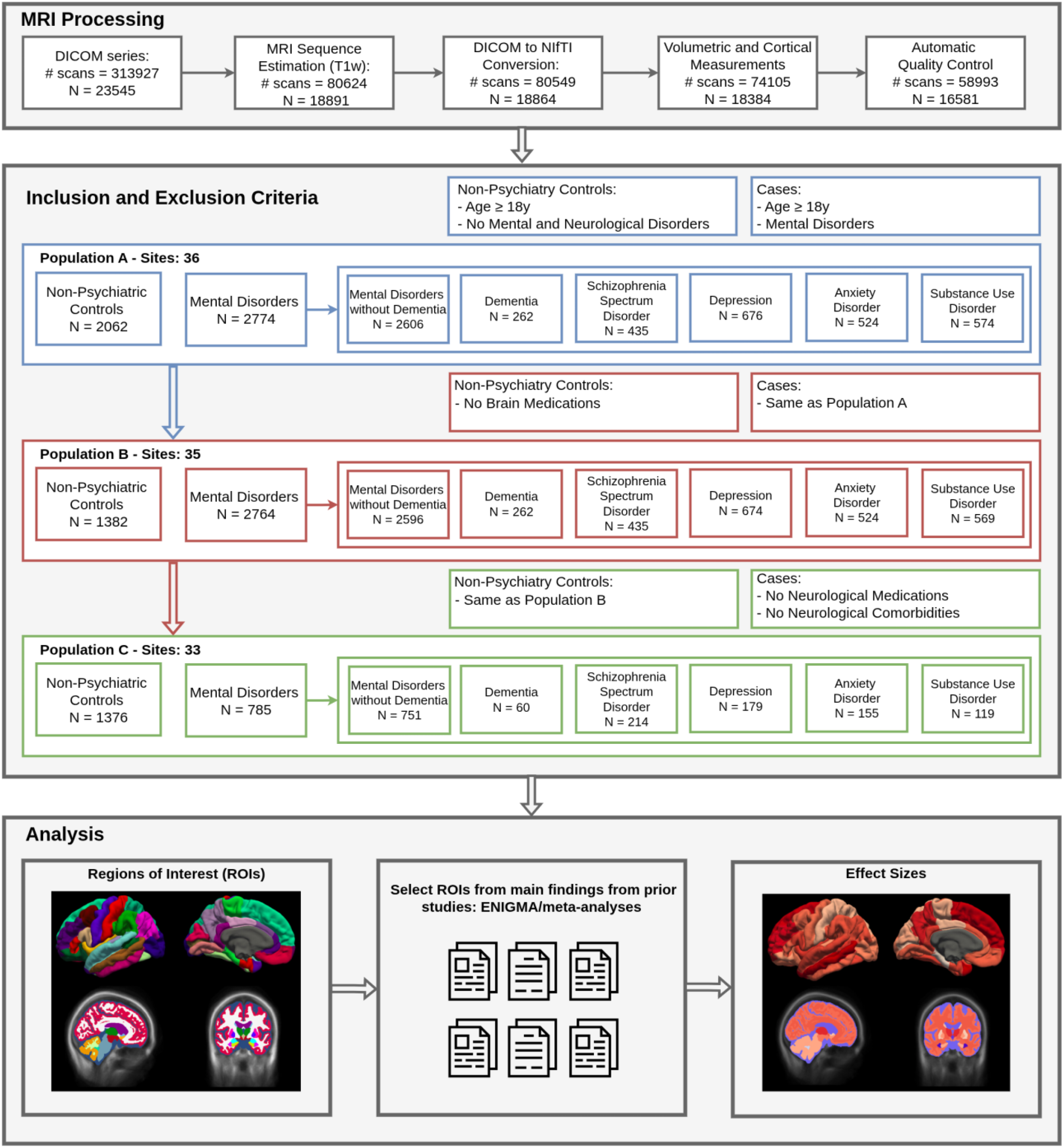
Illustration of the MRI processing pipeline, with selected inclusion and exclusion criteria, and analysis. More details are provided in Supplementary Methods.

### MRI Processing

The analysis focused solely on T1-weighted scans due to their superior contrasts for anatomical structures. We estimated T1-weighted sequences from all available MRI scans, resulting in 80,624 T1-weighted scans (**Supplementary Methods**). Each T1-weighted DICOM sequence was converted to NIfTI using dcm2niix.^22^ We processed each T1w NIfTI scan with recon-all-clinical^11–13,23,24^ to obtain volumetric and cortical thickness estimates. Recon-all-clinical extracts the same cortical and subcortical metrics as the standard FreeSurfer recon-all pipeline^25^ used in ENIGMA studies (**Supplementary Figure S2**), with the addition of extracerebral cerebrospinal fluid volume, and it is optimized for lower-quality, heterogeneous clinical MRI scans. Measurements were averaged bilaterally for regions of interest (ROIs).

All scans were subjected to automated quality control, explicitly designed to remove clear segmentation failures and extreme morphometric outliers while preserving biologically meaningful variability. Low-quality segmentations, as predicted by recon-all-clinical Dice^26^ estimates, were automatically discarded.^11^ Additional quality control involved removing extreme outliers based on interquartile ranges (below or above 2 for each ROI, **Supplementary Methods**). Our quality control steps and inclusion and exclusion criteria allowed us to detect and remove any potential MRI scans affected by severe motion, poor tissue contrast, acquisition-related artifacts, or structural brain abnormalities. Thresholds were computed across the entire population to avoid defining ‘normal’ variation based on any single diagnostic group and to reduce the risk of selectively excluding illness-related morphometric variation. For patients with multiple MRI scans passing quality control, inclusion, and exclusion criteria, the best-predicted scan from recon-all-clinical was used. When performing a sensitivity analysis on slice thickness, we first applied the high- and low-resolution threshold before selecting the best-predicted scan per patient, thereby increasing our sample size and overlap between the two groups. All MRI processing steps are illustrated in **Figure 1** and detailed in **Supplementary Methods**.

### Outcomes of interest

We aimed to replicate known differences between mental disorder patients and non-psychiatric controls, and dementia patients and non-psychiatric controls, based on ROIs identified in previous studies examining regions affected across disorders (10 ROIs: Lateral Ventricles, Thalamus, Amygdala, Hippocampus, Cortical Thickness, Pars Opercularis Thickness, Medial Orbital Frontal Thickness, Rostral Anterior Cingulate Thickness, Superior Temporal Thickness, and Insula Thickness),^3,5–10,16,27^ and well-known studies for dementia (4 ROIs: Lateral Ventricles, Hippocampus, Cortical Thickness, Entorhinal Thickness).^28–30^ For mental disorder subgroups, we identified the following ROIs from previous ENIGMA studies: Substance Use Disorder (7 ROIs: Amygdala, Hippocampus, Insula Thickness, Middle Temporal Thickness, Paracentral Thickness, Precentral Thickness, and Supramarginal Thickness),^9^ Schizophrenia Spectrum Disorder (12 ROIs: Hippocampus, Amygdala, Thalamus, Nucleus Accumbens, Pallidum, Lateral Ventricles, Cortical Thickness, Fusiform Thickness, Inferior Temporal Thickness, Middle Temporal Thickness, Superior Temporal Thickness, and Insula Thickness),^5,6^ Depression (8 ROIs: Hippocampus, Amygdala, Lateral Ventricles, Posterior Cingulate Thickness, Fusiform Thickness, Insula Thickness, Medial Orbitofrontal Thickness, Rostral Anterior Cingulate Thickness),^7,8^ and Anxiety Disorder (4 ROIs: Putamen, Pallidum, Hippocampus, and Amygdala).^10^

### Statistical Analyses

We reported group differences in brain volumes and cortical thickness between cases with mental disorders and non-psychiatric controls for each ROI with multiple linear regression models using statsmodels.^31^ A binary variable, “Diagnosis” (0=non-psychiatric controls, 1=patients), was the predictor of interest. We included sex, age, intracranial volume (ICV) obtained with recon-all-clinical, and site as covariates in the model. Site was defined as the unique combination of scanner and performing hospital; sites without both diagnostic groups were excluded to ensure within-site identifiability of diagnosis effects. The full model is then described as: ROI ~ Diagnosis + Age + Sex +[ICV] + Site, where ICV was included for volumetric measures only. To ease comparisons with prior ENIGMA studies,^5–10^ effect size estimates were calculated using Cohen’s d^32^ computed from the t-statistic of the “Diagnosis” variable from the regression models (Equation 10).^33^ The 95% confidence intervals were derived using Equation 15,^33^ utilizing the asymptotic standard error from Equation 17.^33^ For each comparison, we reported uncorrected two-tailed p-values for the t-statistics of the “diagnosis” binary variable from the linear regression model, accompanied by an “*” in case of significance after multiple comparisons correction for each patient group (Bonferroni, α=0.05).^34^ The number of hypotheses for Bonferroni correction corresponds to the number of ROIs used in each patient group comparison. Differences between our statistical model and those used in prior ENIGMA studies are summarized in Supplementary Table 9. The site distribution for each comparison is provided in Supplementary Tables 10–16.

## RESULTS

In the Capital Region of Denmark, a total of 313,927 MRI images were conducted in 28,129 MRI visits during the year 2019 on 23,545 unique patients. Out of these, a total of 4,836 subjects (2,774 cases and 2,062 non-psychiatric controls) from 36 sites fulfilled the inclusion and exclusion criteria under the most inclusive criteria (Population A), followed by 4,146 subjects from 35 sites for Population B (2,764 cases and 1,382 non-psychiatric controls), and 2,161 subjects from 33 sites for Population C (785 cases and 1,376 non-psychiatric controls) (**Table 1** and **Figure 1**). Most MRI scans were acquired with Philips and Siemens scanners using 3D acquisitions, with magnetic fields of either 1.5T or 3T and slice thicknesses ranging between 1 and 6 mm, with peaks around 1-3 mm. There were no substantial differences in MRI scanner manufacturers, acquisition protocols, or slice thicknesses across criteria (**Supplementary Figure S3**).

**Table 1:** Demographic of the clinical MRI dataset of individuals with mental disorders and non-psychiatric controls.

|  | Population A |  |  |  | Population B |  |  |  | Population C |  |  |  |
| --- | --- | --- | --- | --- | --- | --- | --- | --- | --- | --- | --- | --- |
| Group | N | Sites | Sex [male/female] [female %] | Age [mean ± std] [min, max] | N | Sites | Sex [male/female] [female %] | Age [mean ± std] [min, max] | N | Sites | Sex [male/female] [female %] | Age [mean ± std] [min, max] |
| Mental Disorders (any) | 2774 | 36 | [1589/1185] [43%] | [52.65±18.45] [18, 95] | 2764 | 35 | [1584/1180] [43%] | [52.60±18.46] [18, 95] | 785 | 33 | [458/327] [42%] | [45.56±18.25] [18, 92] |
| Non-Psychiatric Controls | 2062 | 36 | [1217/845] [41%] | [51.37±17.74] [18, 98] | 1382 | 35 | [797/585] [42%] | [48.11±16.93] [18, 98] | 1376 | 33 | [793/583] [42%] | [48.11±16.94][18, 98] |
| Mental Disorders without Dementia | 2606 | 36 | [1509/1097] [42%] | [51.28±18.01] [18, 95] | 2596 | 35 | [1504/1092] [42%] | [51.22±18.01] [18, 95] | 751 | 33 | [438/313] [42%] | [44.30±17.53] [18, 89] |
| Non-Psychiatric Controls | 2062 | 36 | [1217/845] [41%] | [51.37±17.74] [18, 98] | 1382 | 35 | [797/585] [42%] | [48.11±16.93] [18, 98] | 1376 | 33 | [793/583] [42%] | [48.11±16.94][18, 98] |
| Dementia | 262 | 26 | [138/124] [47%] | [75.09±9.82] [34, 93] | 262 | 26 | [138/124] [47%] | [75.09±9.82] [34, 93] | 60 | 16 | [35/25] [42%] | [74.18±9.09] [46, 92] |
| Non-Psychiatric Controls | 1818 | 26 | [1067/751] [41%] | [51.33±17.80] [18, 98] | 1194 | 26 | [689/505] [42%] | [47.75±16.96] [18, 98] | 1048 | 16 | [619/429] [41%] | [48.04±16.84][18, 98] |
| Substance Use Disorder | 574 | 33 | [254/320] [56%] | [58.58±15.06] [19, 90] | 569 | 32 | [252/317] [56%] | [58.50±15.10] [19, 90] | 119 | 20 | [46/73] [61%] | [53.77±14.75] [19, 78] |
| Non-Psychiatric Controls | 2005 | 33 | [1183/822] [41%] | [51.40±17.74] [18, 98] | 1335 | 32 | [772/563] [42%] | [48.12±16.93] [18, 98] | 1201 | 20 | [702/499] [42%] | [47.82±16.88][18, 98] |
| Schizophrenia Spectrum Disorder | 435 | 28 | [213/222] [51%] | [41.29±17.53] [18, 88] | 435 | 28 | [213/222] [51%] | [41.29±17.53] [18, 88] | 214 | 23 | [102/112] [52%] | [36.28±15.97] [18, 78] |
| Non-Psychiatric Controls | 1987 | 28 | [1169/818] [41%] | [51.33±17.74] [18, 98] | 1326 | 28 | [762/564] [43%] | [48.04±16.94] [18, 98] | 1269 | 23 | [734/535] [42%] | [48.00±16.86][18, 98] |
| Depression | 676 | 33 | [459/217] [32%] | [53.28±17.00] [18, 91] | 674 | 32 | [458/216] [32%] | [53.27±17.03] [18, 91] | 179 | 28 | [127/52] [29%] | [48.44±16.57] [18, 85] |
| Non-Psychiatric Controls | 2047 | 33 | [1208/839] [41%] | [51.39±17.73] [18, 98] | 1372 | 32 | [792/580] [42%] | [48.15±16.92] [18, 98] | 1335 | 28 | [774/561] [42%] | [48.04±16.88][18, 98] |
| Anxiety Disorder | 524 | 31 | [365/159] [30%] | [48.06±16.05] [18, 87] | 524 | 31 | [365/159] [30%] | [48.06±16.05] [18, 87] | 155 | 24 | [117/38] [25%] | [43.17±14.88] [18, 84] |
| Non-Psychiatric Controls | 2024 | 31 | [1193/831] [41%] | [51.39±17.76] [18, 98] | 1350 | 31 | [775/575] [43%] | [48.08±16.96] [18, 98] | 1289 | 24 | [743/546] [42%] | [48.14±16.85][18, 98] |
| **Population A:** Non-Psychiatric Controls: age ≥18, No Mental Disorders, No Neurological Disorder. Cases: age ≥18, Mental Disorders.<br>**Population B:** Non-Psychiatric Controls: Population A + No Brain Medications. Cases: Same as Population A.<br>**Population C:** Non-Psychiatric Controls: Same as Population B. Cases: Population A + No Neurological Medications, No Neurological Comorbidities. |  |  |  |  |  |  |  |  |  |  |  |  |

### MRI differences among individuals with Mental Disorders compared to non-psychiatric controls

Results for Population A, B, and C are presented in **Figure 2A-C**, with numerical values in **Supplementary Table S17, Table S18**, and **Table S19**. Under Population C, all comparisons between our clinical MRI data and prior mega-analyses^3,5–10,16,27^ were significant, except for the Rostral Anterior Cingulate after correction for multiple comparisons. Mental disorder patients showed smaller volumes in the thalamus (d=−0.298), amygdala (d=−0.250), and hippocampus (d=−0.145), larger lateral ventricular volume (d=0.272), and a general pattern of thinner cortex (d=−0.180), particularly in the insula (d=−0.177).

**Figure 2:**
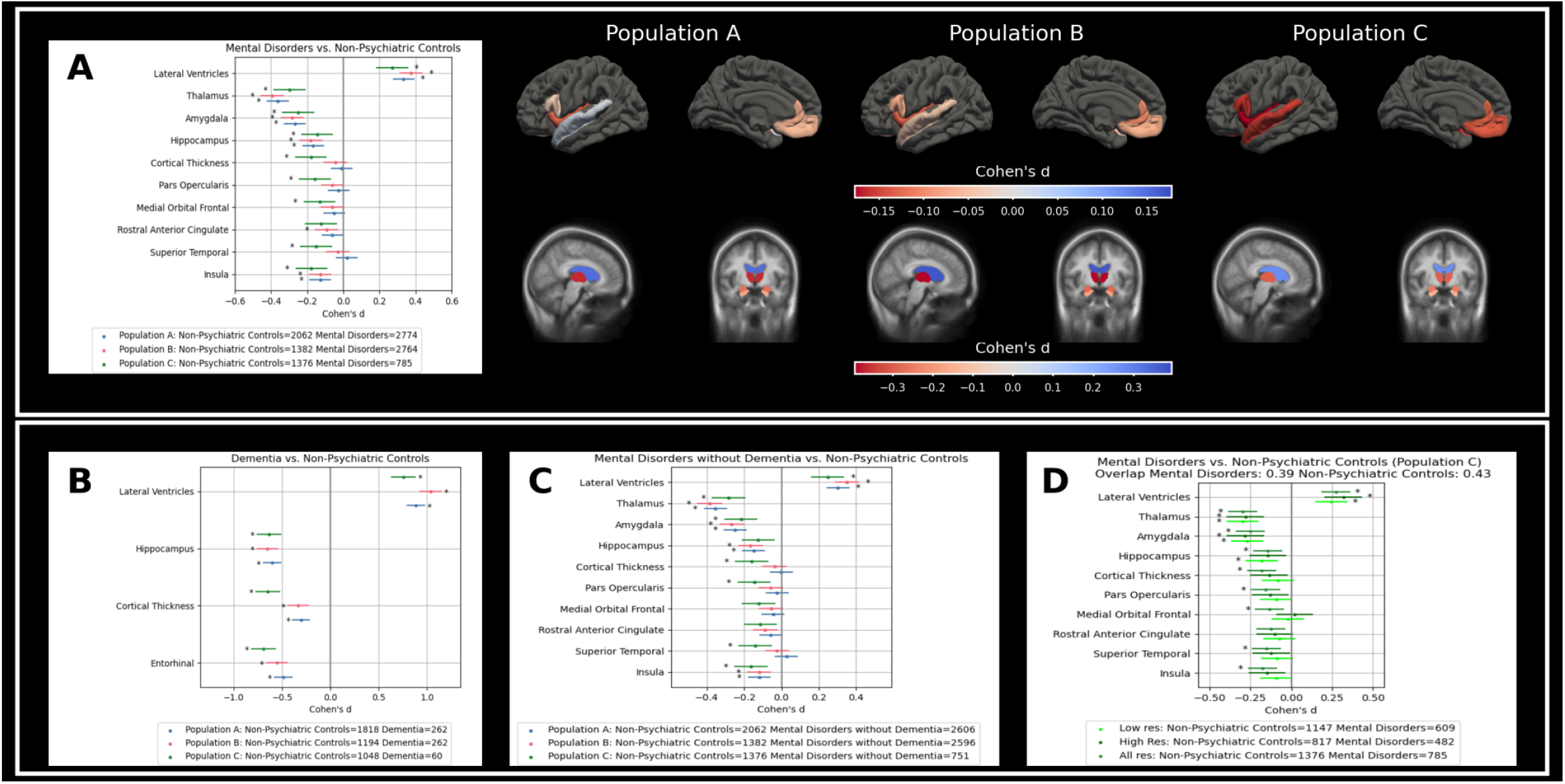
Mental disorders vs. Non-Psychiatric Controls comparisons. **Comparisons between mental disorders and Non-Psychiatric Controls.** Panel A: Patients with mental disorders vs. non-psychiatric controls across inclusion and exclusion criteria. Panel B: Patients with dementia vs. non-psychiatric controls across inclusion and exclusion criteria. Panel C: Patients with mental disorders vs. non-psychiatric controls, excluding dementia patients from the cases group across inclusion and exclusion criteria. Panel D: Impact of slice thickness on differences between patients with mental disorders and non-psychiatric controls for Population C. Results for Population A and B can be found in Supplementary Figure S8. Numerical values for all the figures can be found in Supplementary Table S17 to Table S22.

For cortical thickness, most comparisons under Population A and B showed no significant differences between mental disorder patients and non-psychiatric controls, with a greater pattern of thinner cortex in unmedicated patients and those without brain comorbidities (Population C d=−0.180 vs. Population B d=−0.044). Consistent findings were observed across criteria for volumetric measures, despite Population C having approximately 3.5 times fewer cases and 1.5 times fewer non-psychiatric controls than Population A.

Dementia patients exhibited large effect sizes: smaller hippocampal volume (d=−0.632), larger lateral ventricles (d=0.757), and a general pattern of thinner cortex (d=−0.648), especially in the entorhinal cortex (d=−0.690). To ensure that our findings for mental disorders were not biased by large effect sizes in the dementia subgroup, we performed analysis excluding dementia patients and reassessed Population C (**Figure 2C**). Effect sizes were smaller but consistent with previous findings and all comparisons, except for the hippocampus and Medial Orbitofrontal, were still significant after correction for multiple comparisons.

We identified additional brain differences not reported as main findings in prior ENIGMA studies^3,5–10,16,27^ under Population C (**Figure 3**). After correcting for multiple comparisons across all ROIs, we identified 27 significant differences, highlighting a general pattern of thinner cortex across most cortical areas (cortical thickness d=−0.180, temporal pole d=−0.234, superior frontal d=−0.212, and middle temporal d=−0.191,), smaller deep gray matter structures (thalamus d=−0.298, amygdala d=−0.250, pallidum d=−0.192, and nucleus accumbens d=−0.180), and larger ventricular volumes (inferior lateral ventricles d=0.351, 3rd ventricle d=0.313 and lateral ventricles d=0.272). We also reported higher amounts of extracerebral cerebrospinal fluid (d=0.264). These differences were also found for Population A (**Supplementary Figure S4**) and B (**Supplementary Figure S5**), although to a smaller extent, especially regarding thinner cortex (21 and 22 significant differences, respectively).

**Figure 3:**
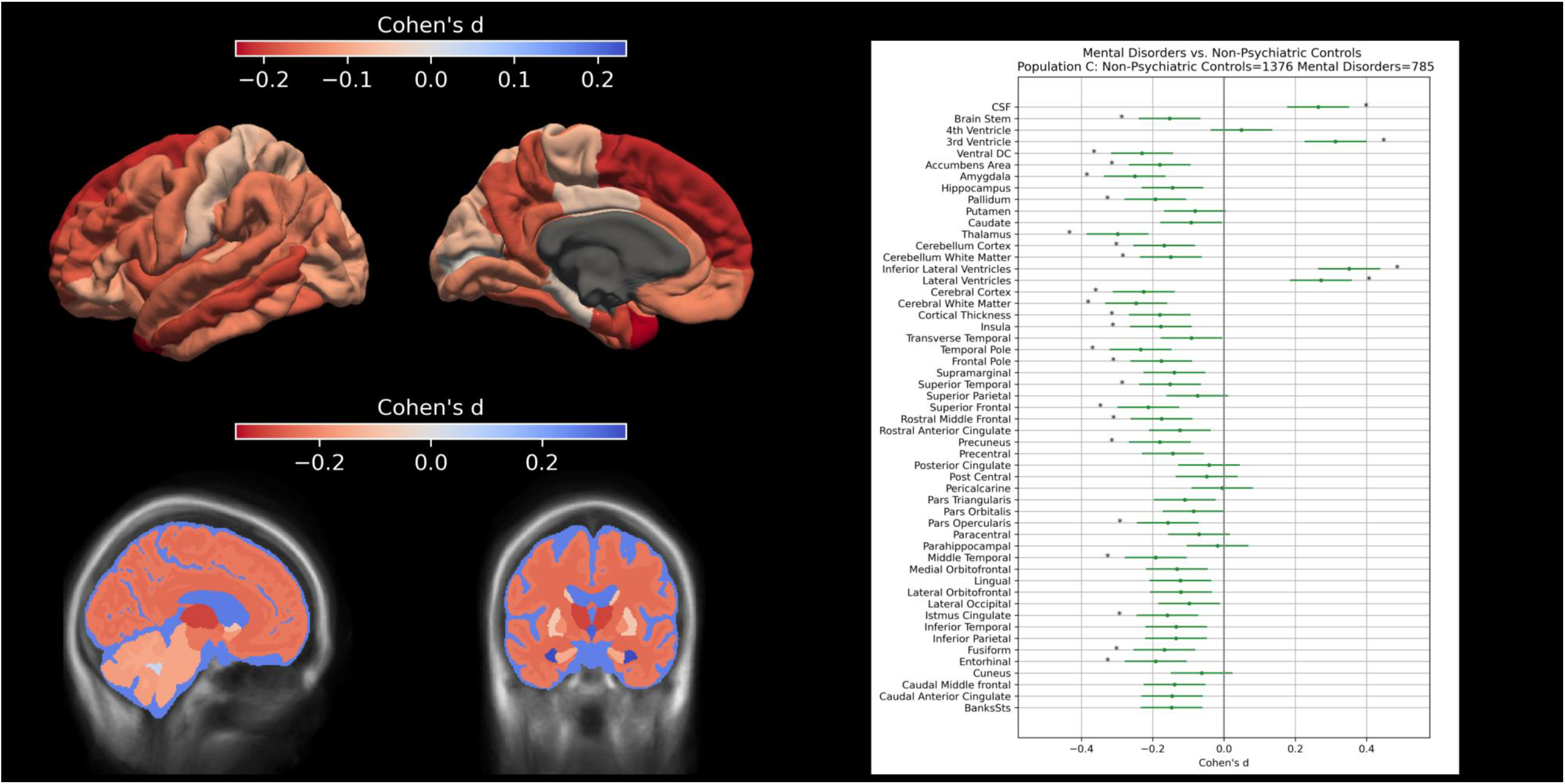
Mental Disorders vs. Non-Psychiatric Controls. Comparisons for all ROI for Population C. Results for Population A and B can be found in Supplementary Figure S4 and Figure S5. Numerical values for all the figures can be found in Supplementary Table S23 to Table S25.

### Comparison between mental disorder subgroups and non-psychiatric controls

For Population B, statistically significant brain differences between mental disorder subgroups and controls were found for 19 of 31 ROIs; and after correction for multiple comparisons, 15 of 31 remained significant (**Figure 4**). Population A and C yielded 14 and 9 significant comparisons, respectively (**Supplementary Figure S6** and **Supplementary Figure S7**). Effect sizes were generally consistent but smaller compared to ENIGMA studies, and most non-significant differences are related to subcortical thickness areas. For the Substance Use Disorder subgroup, we observed larger effect sizes for the amygdala, hippocampus, and insula than previously reported.^9^ Schizophrenia Spectrum Disorder subgroup comparisons revealed consistent patterns of thinner cortex and smaller deep gray matter volumes compared with ENIGMA studies,^5,6^ with only opposite findings for the hippocampus, lateral ventricles, and pallidum. For the Depression subgroup, effect sizes were similar for most comparisons, although mostly not significant, while we noticed a large effect for lateral ventricles compared to ENIGMA.^7,8^ For Anxiety Disorders, we only reported a negative effect for the pallidum in contrast with ENIGMA.^10^

**Figure 4:**
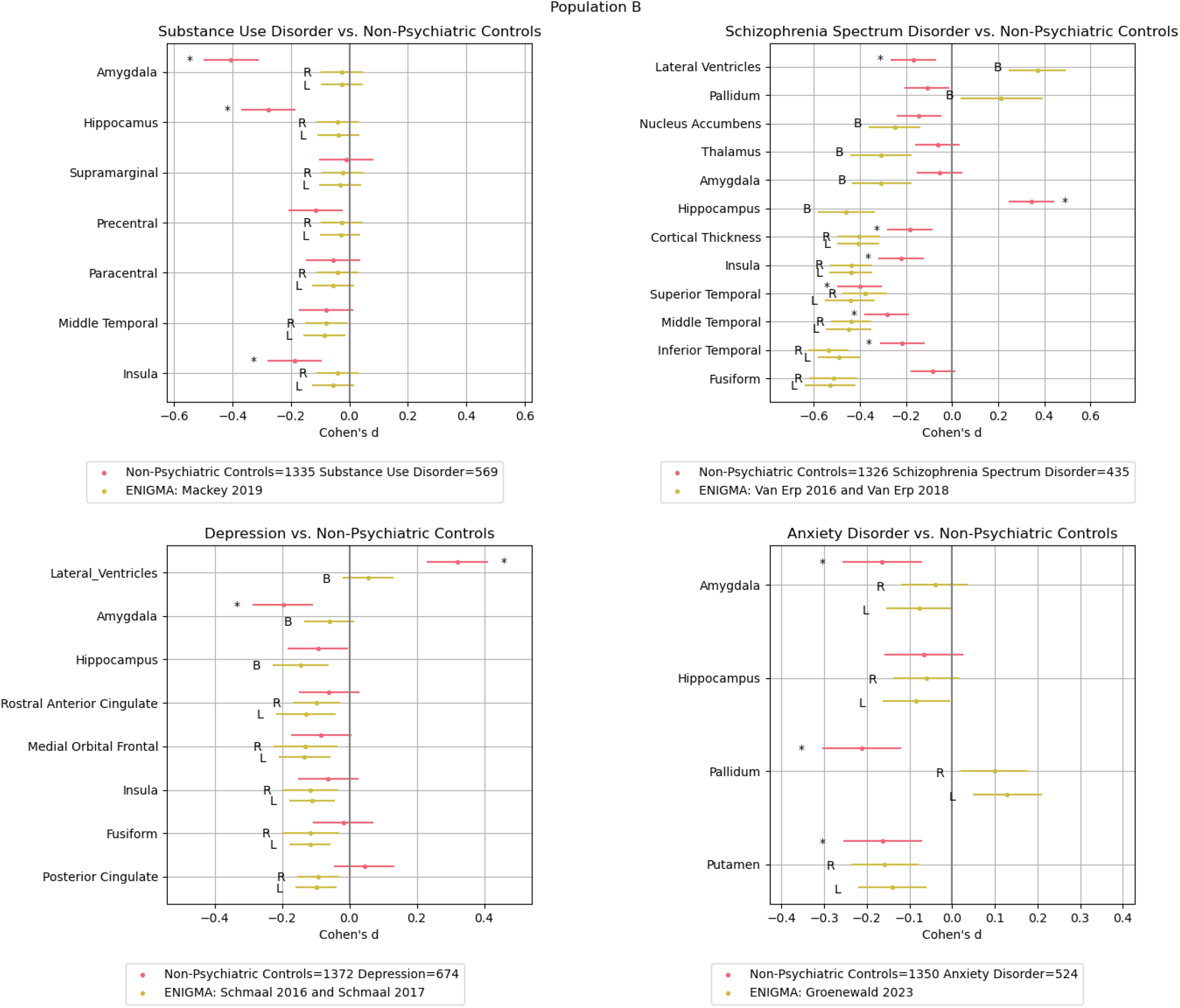
Mental Disorders Subgroups vs. Non-Psychiatric Controls. Comparisons with ENIGMA studies for Population B. R: Right, L: Left, and B: Bilateral. Results for Population A and C can be found in Supplementary Figure S6 and Supplementary Figure S7. Numerical values for all the figures can be found in Supplementary Table S26 to Table S28.

### Effect of Slice Thickness

We assessed the influence of MRI slice thickness on our findings by separating scans into high-resolution (≤1.5 mm) and low-resolution (>1.5 mm) groups (approximately 40% of cases and non-psychiatric controls overlapped between high- and low-resolution groups). Results for Population C are shown in **Figure 2D**. Volumetric comparisons were consistent, while slight differences were observed for cortical thickness comparisons. Similar results were found across criteria for volumetric comparisons, with larger differences in cortical thickness comparisons (**Supplementary Figure S8**).

## DISCUSSION

To our knowledge, this is the first large-scale population-based neuroimaging study using real-world clinical MRI scans to investigate brain changes in mental disorders. We successfully replicated many previously reported findings from meta-analyses of brain differences between patients with mental disorders and non-psychiatric controls. Results in patients with dementia were included as a reference, confirming that the observed differences in other mental disorders were not primarily driven by dementia. However, when examining mental disorder subgroups, we observed smaller effect sizes than those reported in previous meta-analyses. This discrepancy likely stems from differences in the definition of our control group, which included patients undergoing MRI scans for possible brain-related conditions, highlighting future avenues for predicting changes in a more challenging control cohort than typically used in research studies relying on healthy controls. This study is also the largest to date that investigates brain changes associated with mental disorders as a whole, rather than focusing on specific subgroups, as seen in previous ENIGMA studies.^3^ Our findings reveal a general pattern of thinner cortex and smaller deep gray matter volumes among patients with mental disorders broadly consistent with the ENIGMA cross-disorder studies^16–21^, which report overlapping patterns of structural abnormalities across several psychiatric conditions—particularly schizophrenia, bipolar disorder, and major depression—although disorder-specific differences also exist and effect sizes vary considerably across the diagnostic spectrum. These results suggest that clinical MRI data, supplemented with EHR information, can identify biomarkers for mental disorders and improve patient diagnosis and trajectories.

Our exclusion criteria, based on brain medications and comorbidities from EHRs, influenced our findings. We noted consistent effect sizes for volumetric measurements when applying stricter exclusion criteria, even though our sample sizes decreased 3.5-fold for cases and 1.5-fold for non-psychiatric controls. We also observed a more pronounced pattern of thinner cortex in unmedicated patients and those without brain comorbidities compared to the overall patient group. This may reflect selection biases in clinical MRI referrals, whereby unmedicated patients or those without detectable brain comorbidities who nevertheless undergo neuroimaging may represent individuals with more acute or atypical psychiatric presentations, whereas patients with brain comorbidities may be scanned for reasons unrelated to psychiatric disease severity. Furthermore, due to the cross-sectional design, we cannot disentangle medication effects from illness severity or disease duration. Additionally, we found that effect sizes for the Substance Use disorders group were substantially larger for Population C than those reported in a previous ENIGMA study,^9^ as well as our Population A and B, suggesting that different inclusion and exclusion criteria might play an even bigger role, thus warranting further investigation.

When examining mental disorder subgroups, the effect sizes were generally in the same direction as in the ENIGMA studies but often not significant after correcting for multiple comparisons. This might be due to the larger sample sizes in the combined ENIGMA studies, which translate into more significant comparisons, even with small effect sizes. In addition, many ENIGMA cohorts include patients with long-standing illness, whereas our inclusion criteria required a psychiatric diagnosis before or within three months of MRI acquisition. Because MRI is typically performed close to diagnosis in clinical practice, our cohort likely captures individuals at earlier stages of mental disorder disease trajectory with limited cumulative exposure to illness-related processes and treatments. Additionally, our findings suggest that cortical thickness changes are more challenging to detect in clinical MRI scans due to the subtlety of these changes and the lower resolution and quality of clinical images. However, comparing the effect sizes in our study with those reported in ENIGMA studies is complex. Discrepancies in inclusion and exclusion criteria, processing tools, modeling choices, and MRI scan quality between the ENIGMA and our studies likely impact the findings. We used Population B to closely match those in ENIGMA studies. However, several centers in prior ENIGMA studies employed less strict protocols, especially for cases, that are more similar to our less strict criteria (Population A). Indeed, under our strictest criteria (Population C), we observed the least significant comparisons with ENIGMA studies.

The main strength of this study is the use of a large-scale population-based MRI dataset from real-world clinical settings. The integration of EHRs allows for rigorous and flexible inclusion and exclusion criteria, enabling a better understanding of the impact of medication and comorbidities on brain structure, a better match with prior studies’ inclusion and exclusion criteria, and enhancing the ability to track disease progression and correlate brain changes with patient histories.

Limitations include our real-world data with clinical MRI scans being typically of lower quality than those used in research studies, including scans acquired with contrast agents, which may affect tissue contrast and thus segmentation quality. Our findings suggest that differences in effect sizes are only to a small degree influenced by the slice thickness of the scans, particularly for cortical thickness measurements. Furthermore, we could not use established automatic methods like FreeSurfer,^25^ FSL,^35^ and SPM,^36^ as these tools cannot yet handle the low-resolution MRI scans in our clinical cohort. Instead, we employed recently developed automatic tools^11–13^ to obtain cortical and subcortical measurements, which might require further validation. Additionally, we did not perform visual quality control of the segmentations, which is a limitation, as even in ENIGMA studies visual inspection is used to confirm segmentation quality. However, visual quality control is often infeasible in large-scale clinical datasets. Although site effects were modeled by scanner and hospital in our linear model, we did not apply more complex harmonization techniques like ComBat^37^ because several sites included very small sample sizes, which can yield unstable estimates. A prior ENIGMA study^38^ has shown that effect sizes can vary depending on site-effect modeling strategies, highlighting the importance of evaluating harmonization approaches when sufficient site-specific data are available.

Our cohort is the largest population-based cohort, yet smaller than the combined worldwide cohorts in prior ENIGMA mega-analyses studies. However, our study is based on MRI scans acquired in a single year (2019) only, thus, there is substantial potential in future studies to mitigate the subtle nature of subcortical differences, where more data is needed to validate our findings further. Moreover, we did not age- and sex-match controls in this study due to the limited sample size, but we adjusted for these covariates in our analysis. Our sample size was also yet insufficient for studying brain changes in other mental disorders such as bipolar disorder, attention-deficit/hyperactivity disorder, and autism spectrum disorder. Additionally, future work may investigate how factors such as duration of illness, age at onset, cumulative medication exposure, and sociodemographic factors may influence cortical thickness and volumetric changes in mental disorders.

Clinical MRI scans can effectively detect cortical and subcortical differences among patients with mental disorders and also distinguish them from other patients without mental disorders. When examining mental disorder subgroups, we observed consistent, though smaller, effect sizes compared to previous ENIGMA mega-analyses studies using high-quality MRI data. These differences are likely due to our control group comprising patients undergoing MRI scans for potential clinical diagnoses. In contrast, the studies in the ENIGMA mega-analyses compare to healthy controls, which makes prediction easier, whereas, in a real-world clinical context, the separation from other individuals undergoing examination is of importance for specificity.

Our findings reveal shared brain differences across mental disorders. Factors such as slice thickness, comorbidities, and medication can affect the detection of structural changes. Population selection should reflect the study objective: real-world clinical samples (Population A) capture brain patterns representative of routine care but introduce greater heterogeneity, whereas strictly defined samples (Population C) minimize variability and improve specificity, at the cost of broader generalizability. We recommend standardizing inclusion/exclusion criteria, incorporating EHR-based comorbidity data, and leveraging routine clinical MRI scans—including lower-resolution images—to enable large-scale biomarker discovery while accounting for scan quality. These strategies can support more precise investigations of neurobiological mechanisms and advance diagnostic and therapeutic approaches.

## Data Availability

All data is available to all researchers based in Denmark and employed in the Capital Region of Denmark after the relevant approvals from the Danish health authorities

## ACKNOWLEDGEMENTS

Stefano Cerri has received funding from the Lundbeck Foundation (R449-2023-1512). Mostafa Mehdipour Ghazi has received funding from the Lundbeck Foundation (R400-2022-617). Mads Nielsen is supported by the Pioneer Centre for AI, Danish National Research Foundation, grant number P1. Michael Eriksen Benros has received funding from the Lundbeck Foundation (R278-2018-1411 and R383-2022-285).

