## Supplementary Material for "Cross-disorder comparison of Brain Structures among 4,836 Individuals with Mental Disorders and Controls utilizing Danish population-based Clinical MRI Scans"

**Supplementary Methods.** MRI processing pipeline, inclusion, and exclusion criteria.
**Supplementary Figure S1.** Sensitivity analysis on the number of unique patients who received a diagnosis either before or after the MRI scan was taken, measured in months, and included patients who were diagnosed within 3 months before or after the date of the MRI.
**Supplementary Figure S2.** Regions of interest.
**Supplementary Figure S3.** MRI dataset information.
**Supplementary Figure S4.** Comparisons of all ROIs for Population A.
**Supplementary Figure S5.** Comparisons of all ROIs for Population B.
**Supplementary Figure S6.** Comparison of findings between Mental Disorders Subgroups and Non-Psychiatric Controls, with prior ENIGMA studies for Population A.
**Supplementary Figure S7.** Comparison of findings between Mental Disorders Subgroups and Non-Psychiatric Controls, with prior ENIGMA studies for Population C.
**Supplementary Figure S8.** Comparison of findings for high- (≤1.5mm slice thickness) and low-resolution (>1.5mm slice thickness) scans for Population A and Population B.
**Supplementary Table S9.** Comparison of statistical models used in ENIGMA studies and the present study.
**Supplementary Table S10.** Site distribution of Mental Disorders (any) patients and Non-Psychiatric Controls.
**Supplementary Table S11.** Site distribution of Mental Disorders without Dementia and Non-Psychiatric Controls.
**Supplementary Table S12**. Site distribution of Dementia patients and Non-Psychiatric Controls.
**Supplementary Table S13.** Site distribution of Substance Use Disorder patients and Non-Psychiatric Controls.
**Supplementary Table S14.** Site distribution of Schizophrenia Spectrum Disorder patients and Non-Psychiatric Controls.
**Supplementary Table S15.** Site distribution of Depression patients and Non-Psychiatric Controls.
**Supplementary Table S16.** Site distribution of Anxiety Disorder patients and Non-Psychiatric Controls.
**Supplementary Table S17.** Comparison of Mental Disorders and Dementia patients for previously reported ROI differences for Population A.
**Supplementary Table S18.** Comparison of Mental Disorders and Dementia patients for previously reported ROI differences for Population B.
**Supplementary Table S19.** Comparison of Mental Disorders and Dementia patients for previously reported ROI differences for Population C.
**Supplementary Table S20.** Comparison of findings for high- (≤1.5mm slice thickness) and low-resolution (>1.5mm slice thickness) scans for Population A.
**Supplementary Table S21.** Comparison of findings for high- (≤1.5mm slice thickness) and low-resolution (>1.5mm slice thickness) scans for Population B.
**Supplementary Table S22.** Comparison of findings for high- (≤1.5mm slice thickness) and low-resolution (>1.5mm slice thickness) scans for Population C.
**Supplementary Table S23.** Comparisons of all ROIs for Population A.
**Supplementary Table S24.** Comparisons of all ROIs for Population B.
**Supplementary Table S25.** Comparisons of all ROIs for Population C.
**Supplementary Table S26.** Comparison of findings between Mental Disorders Subgroups and Non-Psychiatric Controls, with prior ENIGMA studies for Population A.
**Supplementary Table S27.** Comparison of findings between Mental Disorders Subgroups and Non-Psychiatric Controls, with prior ENIGMA studies for Population B.
**Supplementary Table S28.** Comparison of findings between Mental Disorders Subgroups and Non-Psychiatric Controls, with prior ENIGMA studies for Population C.

### Supplementary Methods: MRI processing pipeline, inclusion, and exclusion criteria

This section describes how we processed the clinical MRI dataset and selected the inclusion and exclusion criteria.

#### 1) MRI sequence estimation

We assigned every scan to one or multiple of the following categories: Fluid-attenuated inversion recovery (FLAIR), Diffusion- (DWI), Susceptibility- (SWI), T1- (T1), T2-weighted imaging (T2* excluded), as well as other identified sequences. The categorization was performed by checking the DICOM header entry Series Description (0008,103E) for the following substrings:

- T1-weighted imaging: 'T1', 't1', 'BRAVO', 'mprage', 'MPRAGE' (49,108 scans);
- T2-weighted imaging: 'T2', 't2' (47,679 scans);
- Fluid-attenuated inversion recovery: 'FLAIR', 'flair', 'Flair' (21,322 scans);
- Susceptibility-weighted imaging: 'SWI', 'swi', 'SUSCEPTABILITET' (20,434 scans);
- Diffusion-weighted Imaging: 'DWI', 'dwi', 'MUSE', 'Diffusion', 'DTI', 'dti', (35,521 scans);
- Other identified sequence types: 'vessel scout', 'VRT', 'csf flow', 'WIP', 'svs', 'SVS', 'animation', 'ACOM', 'Verification', 'BA', 'BLACK BLOOD', 'Batch', 'Cerebral Blood Flow', 'DISPLAY', 'IMPAX Volume', 'MYELO', 'MI Reading', 'MM Oncology Reading', 'SCREENSAVE', 'SPINE', '3D Saved State - AutoSave', 'Screen Save', 'MINIP', 'Minimum Intensity', 'Min Intensity Projection', 'Min Intensity ', 'Minimum Intensity Projection', 'Tra SWIp MinIP', 'BOLD', 'Loc', 'loc', 'Scout', 'LOC', 'lokal', 'LOKAL', 'B1 Calibration', 'calib', 'Calib', 'cal', 'Cal', 'CAL', 'PDW', 'TOF', 'ToF', 'tof', 'angio', 'Angio', 'ANGIO', 'SWAN', 'PCA', 'pca', 'dce', 'PC', 'pc', 'TRANCE', 'trance', 'mIP', 'MIP', 'SURVEY', 'Survey', 'survey', 'CEST', 'ASL', 'asl', 'TRACEW', 'travew', 'STIR', 'stir', 'ADC', 'Apparent Diffusion Coefficient', 'adc', 'Perfusion Weighted', 'T2*', 't2*' (71,814 scans).

Based on the categorization, the second step was constructing classes used as training and prediction targets. Scans from the T2 category are assigned to the T2 class except those in the FLAIR category, which were then assigned to the FLAIR class. The intersection of FLAIR and T2 categories occurs because FLAIR is a T2-weighted MRI technique modification often called "T2-FLAIR". Scans in other intersections of categories were assigned to the "Unlabeled" class. We predicted a label (FLAIR, DWI, SWI, T1, T2, or other identified sequence) for every unlabeled scan (N=71,049), after training and validation on the labeled data (N=242,878). As input features, we used non-private DICOM header tags that might be related to the sequence type. Tags with more than half the entries missing are discarded. The remaining tags used for classification are:

- (0018,0081) Echo Time;
- (0018,0080) Repetition Time;
- (0018,1314) Flip Angle;
- (0018,0091) Echo Train Length;
- (0018,0020) Scanning Sequence;
- (0018,0021) Sequence Variant;
- (0018,1318) dBdt;
- (0018,0022) Scan Options;
- (0018,0084) Imaging Frequency;
- (0018,0086) Echo Numbers.

Missing numeric values were replaced by the median of the feature computed over all scans. The scanning sequence (SS), sequence variant (SV) and, scan options (SO), image type (IT) features were one-hot encoded, i.e., every distinct possible entry (e.g., Spin Echo, Inversion Recovery, Magnetization Transfer Contrast, etc) is represented by a binary vector. Entries that occurred in less than 1% of all training samples were dropped. Some entries occurred under multiple tags, e.g., Inversion Recovery was present under the Scan Options and the

Scanning Sequence tag. These were combined with an OR statement to one binary feature. For SV, SS, and SO tags, empty lists were also converted to binary features. The preprocessing results in 74 features (67 binary, 6 continuous, and 1 discrete). Labeled data is split into a train (N=218,590) and a validation set (N=24,288). XGBoost (xgboost, v.2.0.3)[1] was used for classification. To find good hyperparameters, 5-fold cross-validation was performed using Bayesian optimization. For more robust classification performances, tuning and training of the classifier were repeated 5 times with different initializations of the model's parameter, and majority voting was performed to obtain the final classification. In the case of ties, the T1w class was picked. After sequence prediction, we obtained 80,624 T1w, 51,685 T2w, 24,290 FLAIR, 23,406 SWI, 34,383 DWI, and 99,539 "other" scans.

**2) MRI Dicom to NIfTI conversion**

We converted to NIfTI all the 80,624 T1w DICOM series obtained after the MRI sequence estimation with dcm2niix (v.1.0.20230411)[2]. 75 conversions resulted in an error and the corresponding scans were discarded. When multiple NIfTI scans were obtained from a single DICOM series, the largest converted file was kept for further processing.

**3) Recon-all-clinical**

We processed each of the 80,549 converted NIfTI T1w scans with recon-all-clinical[3-6] (FreeSurfer v. 7.4.1). 6,444 scans resulted in errors; this is due to a limited field of view of the images, 2D images, or MRI pathologies that result in an error at any step of the pipeline.

**4) Automatic Quality Control**

Recon-all-clinical provides estimated Dice[7] scores for the following brain structures:[5] white matter, cortex, lateral ventricle, cerebellum, thalamus, hippocampus, amygdala, pallidum, putamen, and brainstem. We considered a segmentation failure when the Dice score of any structure listed above was below 0.65[5]. 11,549 scans were discarded.

All regions of interest (ROIs) of Sections 3.2 and 3.3, normalized for intracranial volume, with a volume or thickness larger or smaller than 2 times the interquartile range were discarded for further automatic quality control. 3,563 scans were discarded. 58,993 scans remained after automatic quality control.

Age and sex distributions were preserved before and after interquartile range–based filtering (mean age: 55.24 vs. 55.23 years; proportion female: 57.0% vs. 57.2%). After applying inclusion and exclusion criteria (defined below), interquartile range–based filtering in Population A reduced the sample from 9,659 mental disorder patient scans to 9,060 scans (6.2% excluded) and from 7,204 non-psychiatric control scans to 6,984 scans (3.1% excluded). The higher exclusion rate for patient scans reflects that multiple scans were often acquired per MRI visit, typically to replace earlier scans of insufficient quality.

**5) Inclusion and exclusion criteria**

We have the following three populations based on different criteria for inclusion and exclusion. Diagnostic and medication codes were based on ICD-10 Version: 2019[8]:

**Population A:**

***Non-psychiatric Controls:***
For non-psychiatric controls, the following criteria were used:

- No psychiatric diagnosis (F00-F99, G30) before or within 3 months after the acquisition of the MRI scan;
- age >= 18 years at the time of the MRI scan;

- No neurological comorbidities:
    - Nervous system: G00-G99, except G560 (Carpal tunnel syndrome) and G562 (Ulnar nerve neuropathy);
    - Cerebrovascular diagnosis: I60-69, E236E;
    - CNS infections: A022C, A066, A17, A229C, A321, A390, A504, A514B, A521A–B, A548A, A80–A89, B003–B004, B010–B011, B020–B021, B050–B051, B060, B261–B262, B375, B451, B582, B602, E236A;
    - Traumatic brain injury: S060-S069;
    - Skull fracture: S020, S021, S027, S029;
    - Malignant cancer: C00-C99;
    - Benign neoplasm in the nervous system: D32-D33;
    - HIV: B20-B24.

***Cases with mental disorders***:
For patients with mental disorders, the following diagnoses were considered if present in the electronic health record:

- Psychiatric diagnosis: F00-F99, G30
- diagnosis before or within 3 months after the acquisition of the MRI scan;
- age >= 18 years at the time of the MRI scan;

**Population B:**

***Non-psychiatric Controls:***
Further exclusion criteria on Population A based on medication history. Specifically, we removed controls having brain medication utilizing the codes starting with N0[3-7], except for N05AD08 (Droperidol, anti-nausea medication) and N07BA01 (Nicotine, aid in quitting smoking).

***Cases with mental disorders***: Same as Population A.

**Population C:**

***Non-psychiatric Controls***: Same as Population B.

***Cases with mental disorders***:
Further exclusion criteria on Population B based on neurological medication history and neurological comorbidities:

- brain medication starting with N04.
- No neurological comorbidities:
    - Nervous system: G00-G99, except G560 (Carpal tunnel syndrome) and G562 (Ulnar nerve neuropathy);
    - Cerebrovascular diagnosis: I60-69, E236E;
    - CNS infections: A022C, A066, A17, A229C, A321, A390, A504, A514B, A521A–B, A548A, A80–A89, B003–B004, B010–B011, B020–B021, B050–B051, B060, B261–B262, B375, B451, B582, B602, E236A;
    - Traumatic brain injury: S060-S069;
    - Skull fracture: S020, S021, S027, S029;
    - Malignant cancer: C00-C99;
    - Benign neoplasm in the nervous system: D32-D33;
    - HIV: B20-B24.

**Supplementary Figure S1: Sensitivity analysis on the number of unique patients who received a diagnosis either before or after the MRI scan was taken, measured in months, and included patients who were diagnosed within 3 months before or after the date of the MRI** (The results are based on the most inclusive criteria (Population A)).

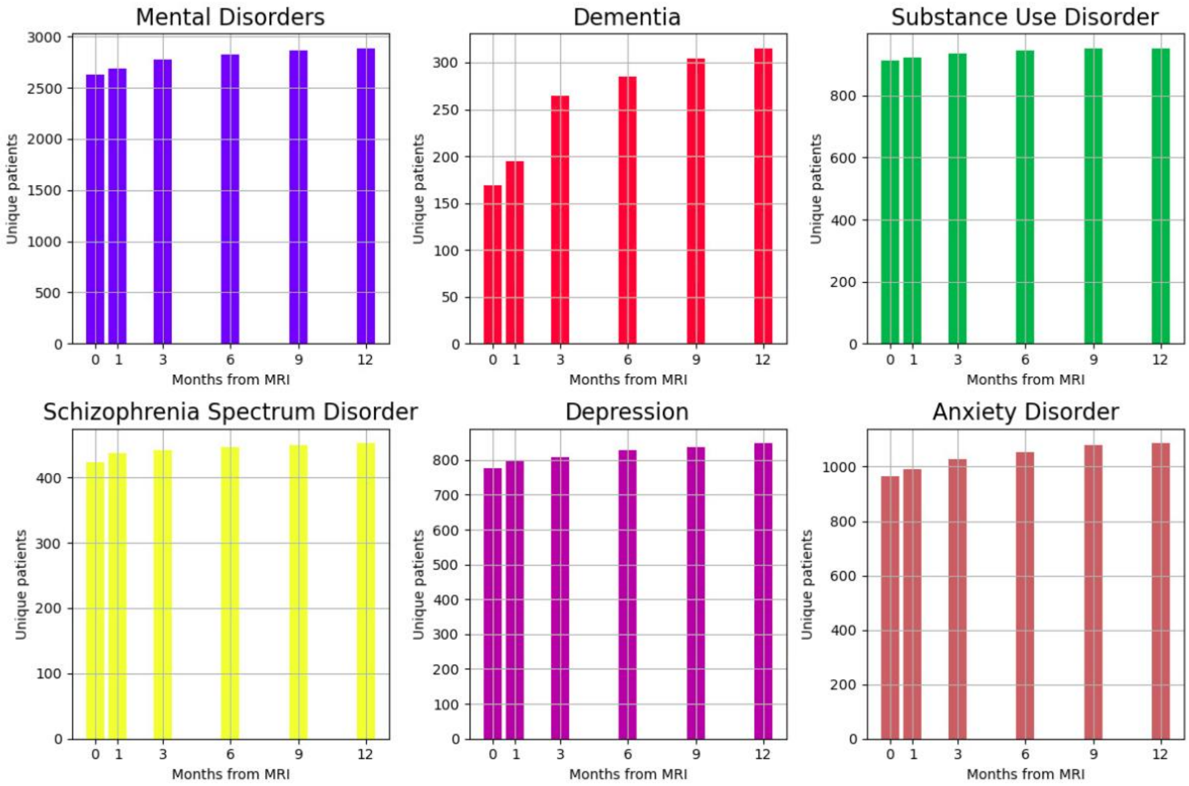

**Supplementary Figure S2: Regions of interest. DK atlas parcellation[9] and recon-all-clinical whole-brain segmentation on "fsaverage" from FreeSurfer[10].**

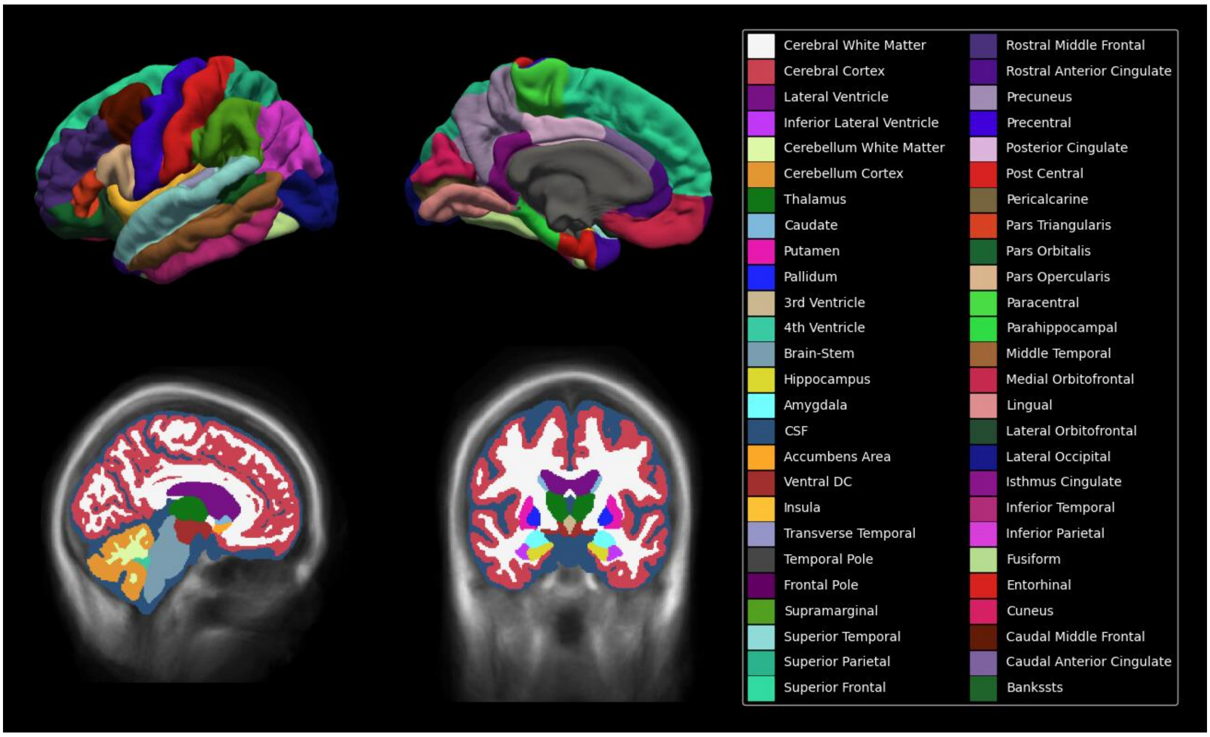

**Supplementary Figure S3: MRI dataset information.**

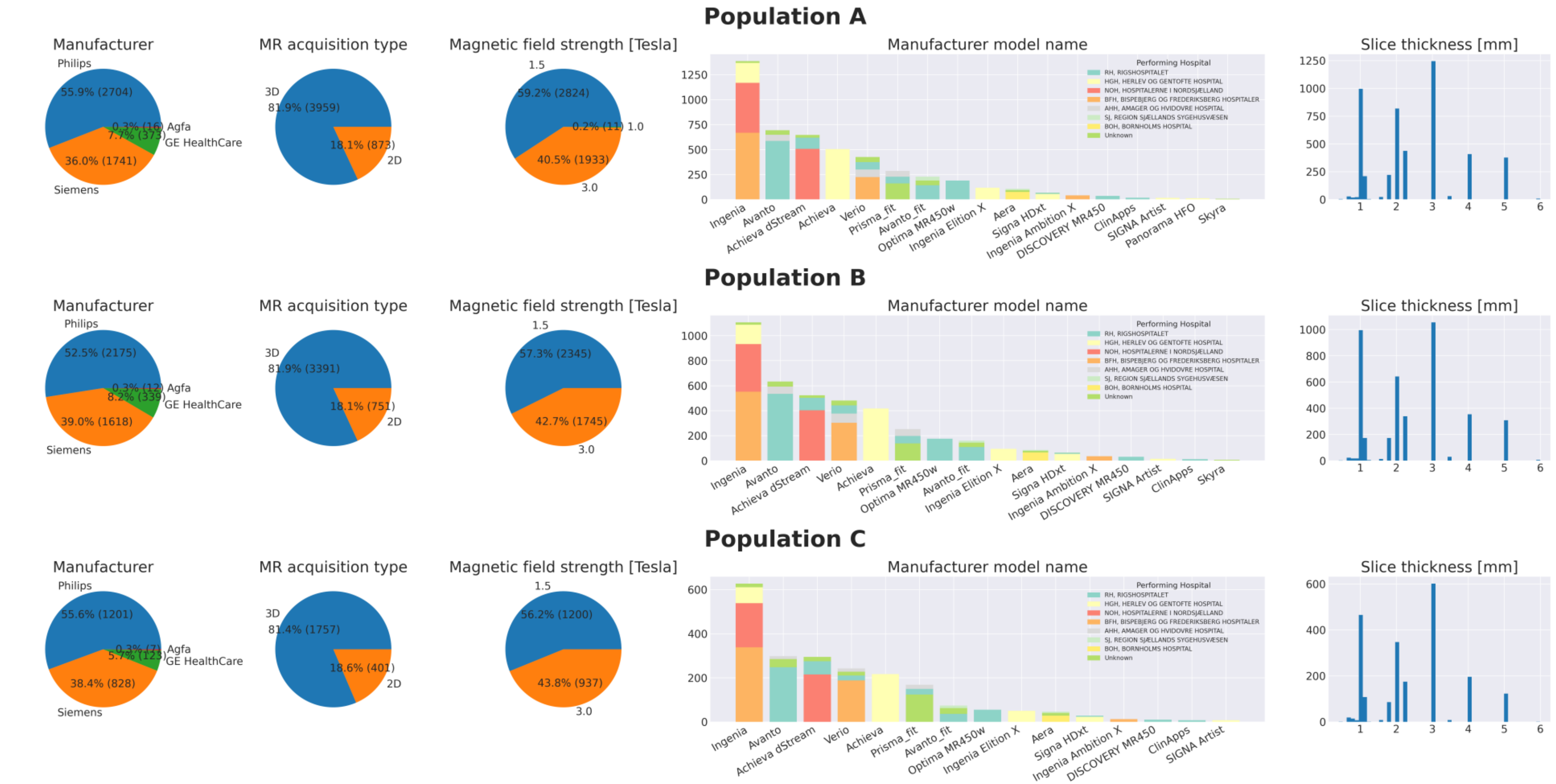

**Supplementary Figure S4: Comparisons of all ROIs for Population A.** Numerical values are provided in Supplementary Table S23.

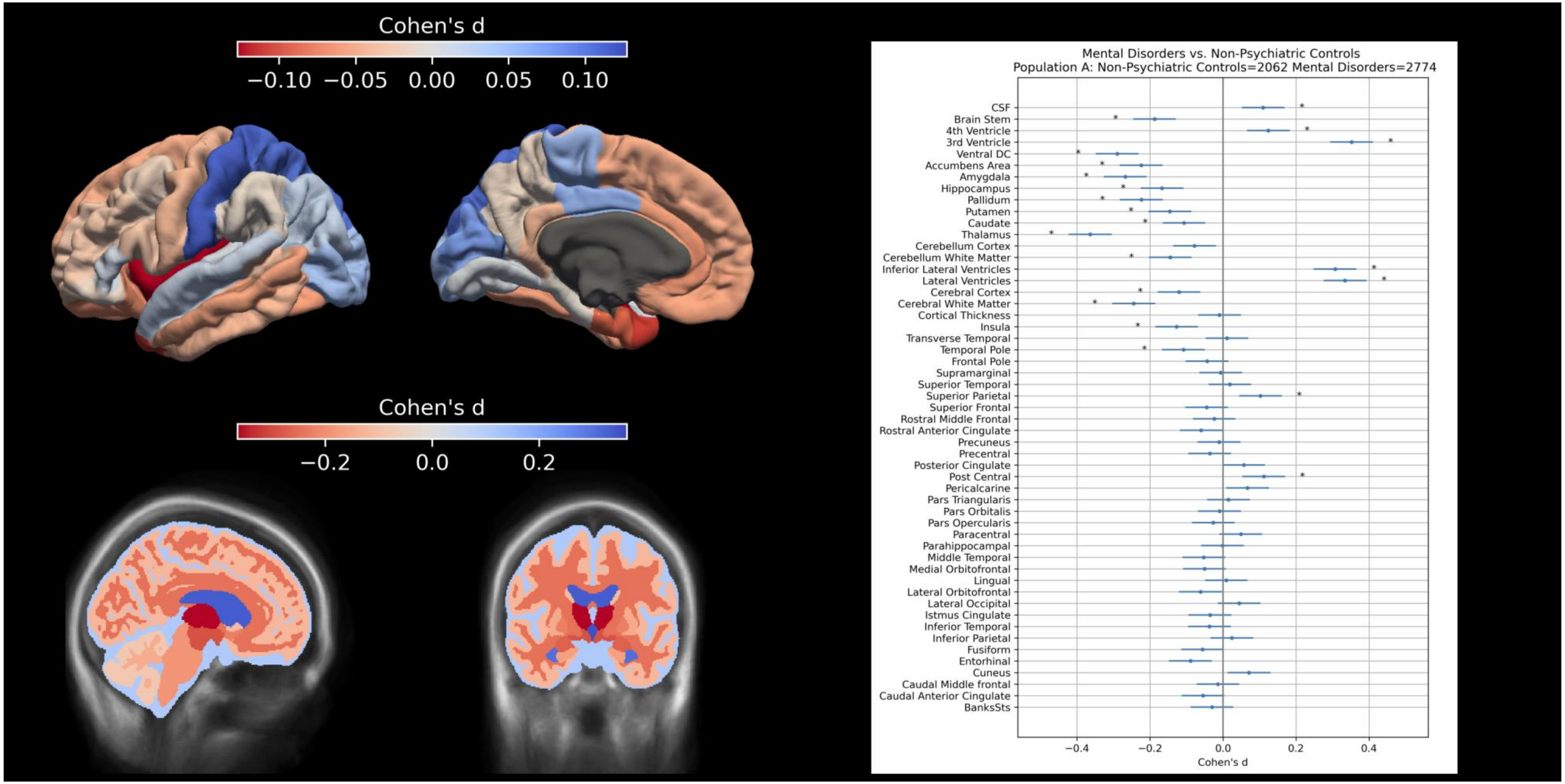

**Supplementary Figure S5: Comparisons of all ROIs for Population B.** Numerical values are provided in Supplementary Table S24.

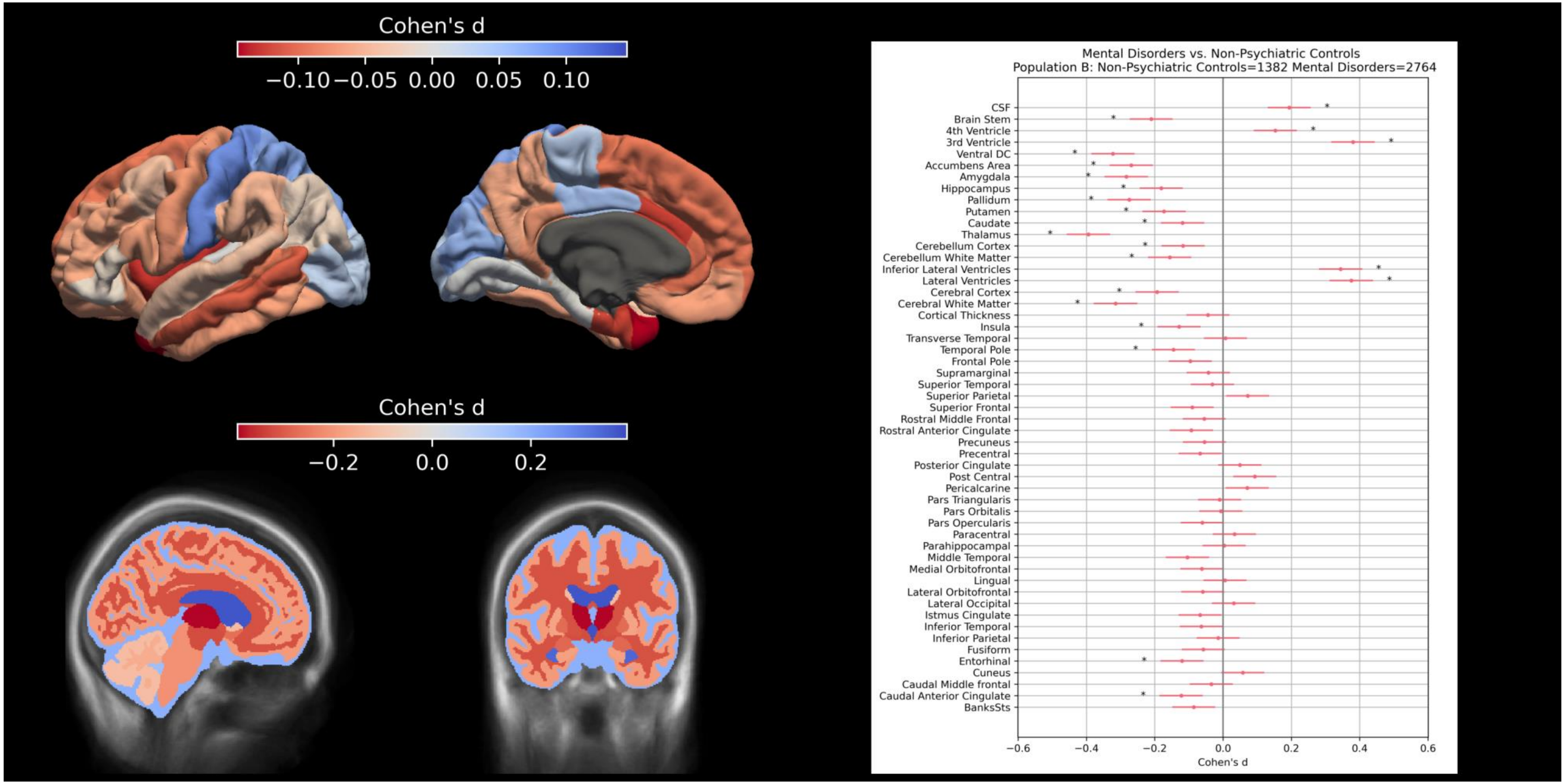

**Supplementary Figure S6: Comparison of findings between Mental Disorders Subgroups and Non-Psychiatric Controls, with prior ENIGMA studies for Population A.** R=Right hemisphere, L=Left hemisphere, B=Bilateral. Numerical values are provided in Supplementary Table S26.

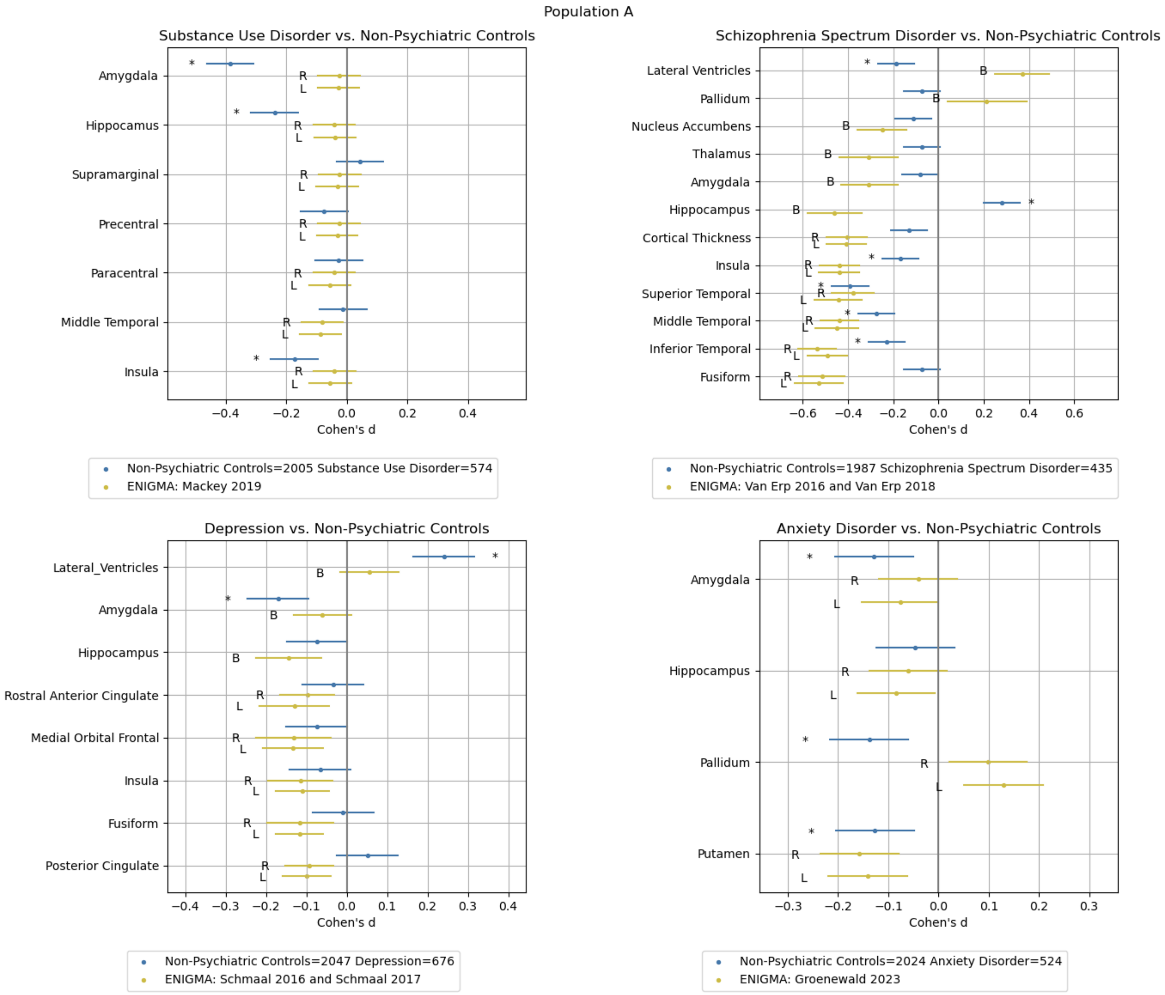

**Supplementary Figure S7: Comparison of findings between Mental Disorders Subgroups and Non-Psychiatric Controls, with prior ENIGMA studies for Population C.** R=Right hemisphere, L=Left hemisphere, B=Bilateral. Numerical values are provided in Supplementary Table S28.

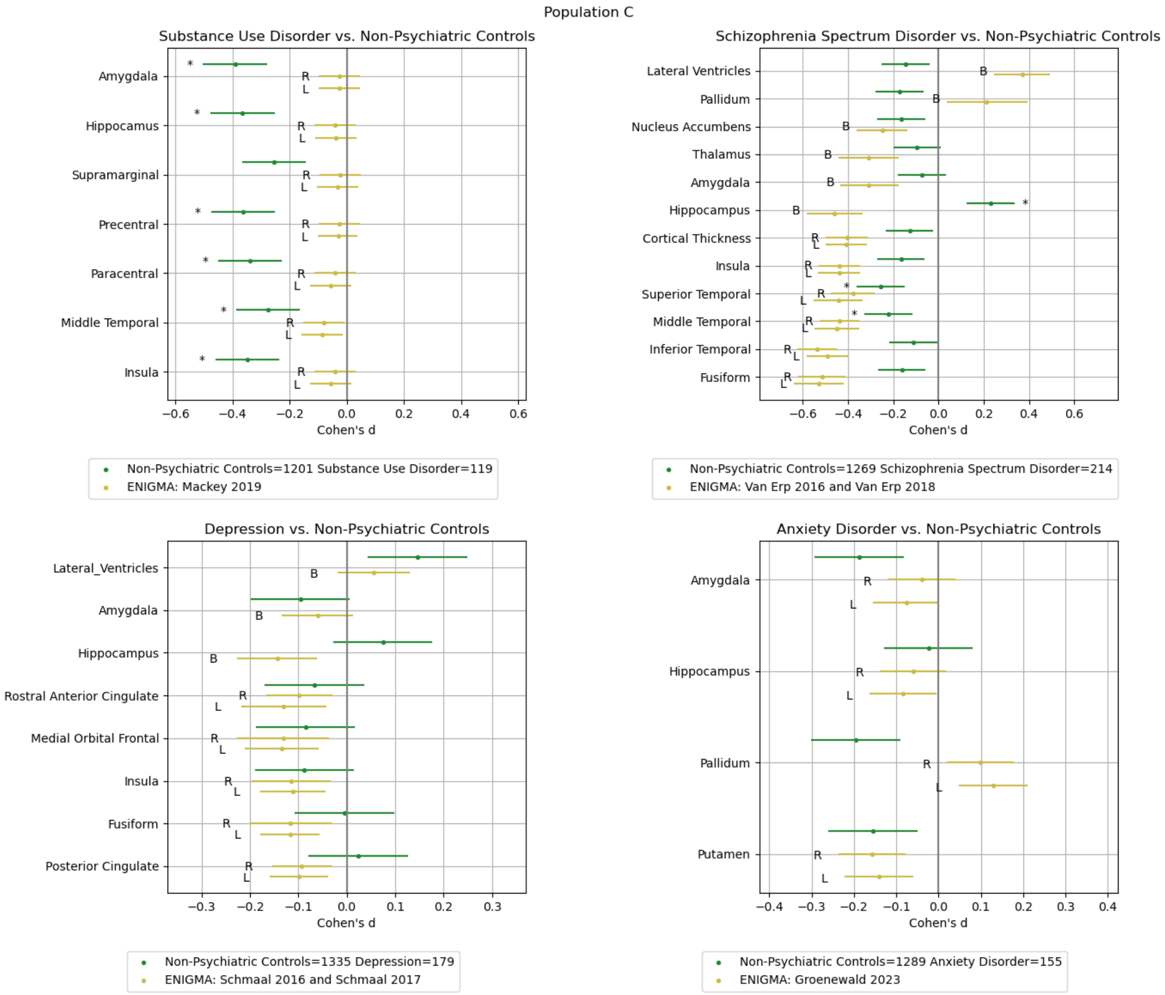

**Supplementary Figure S8: Comparison of findings for high- (≤1.5mm slice thickness) and low-resolution (>1.5mm slice thickness) scans for Population A and Population B.** Numerical values are provided in Supplementary Table S20 and Table S21.

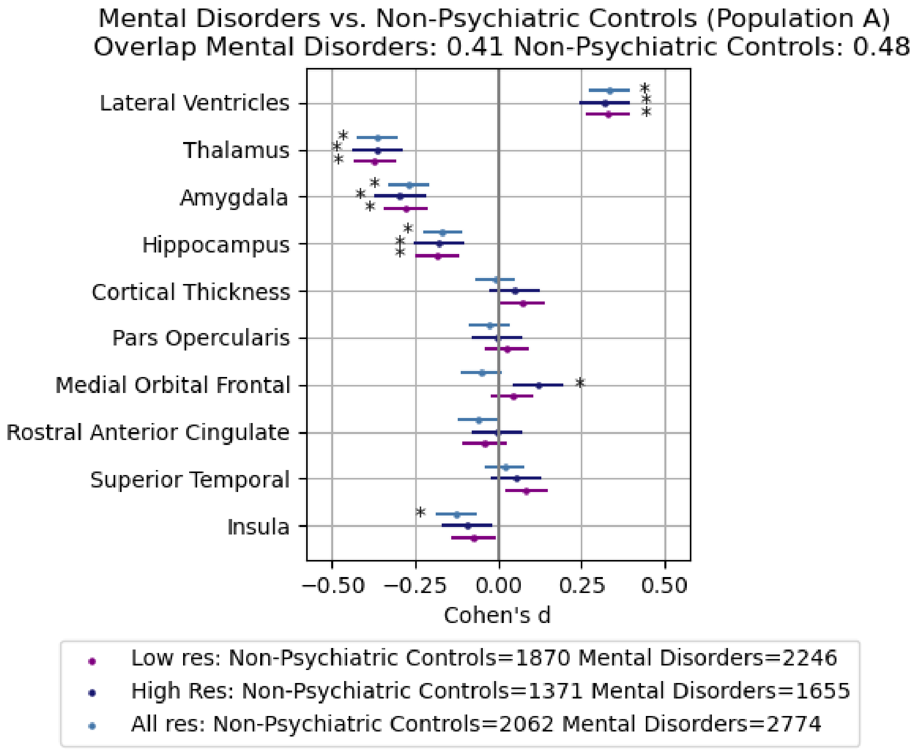


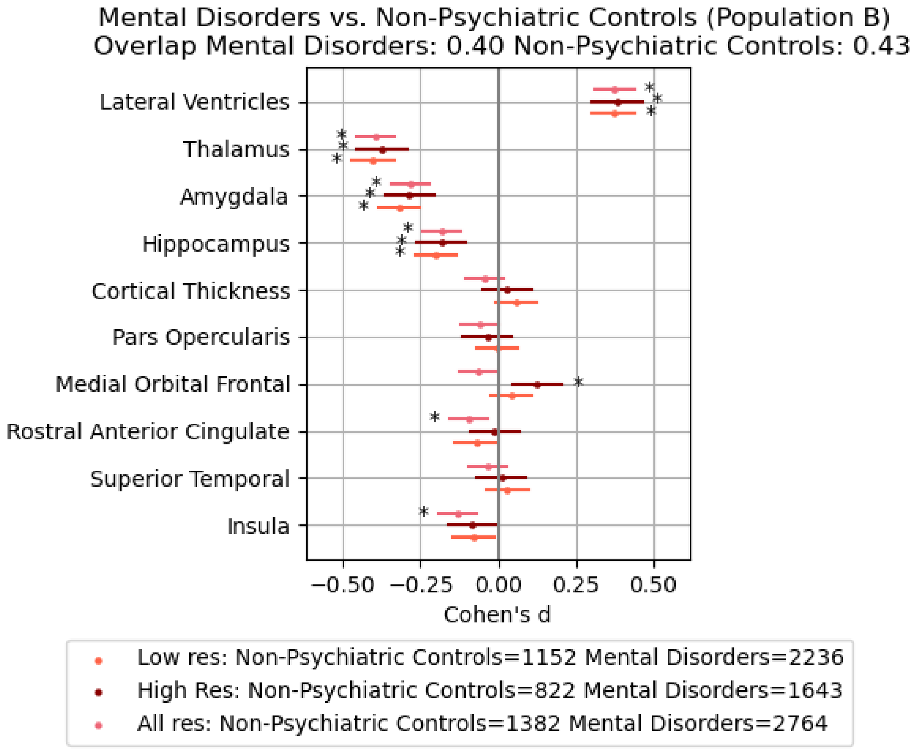

**Supplementary Table S9. Comparison of statistical models used in ENIGMA studies and the present study.**

| ENIGMA study | Model | ROI | Multiple comparisons | Notes |
|---|---|---|---|---|
| Mackey 2019 [11] | ROI ~ Diagnosis+ Sex + Age + ICV + (1\|Site) | Volumes and thicknesses | False discovery rate | Mixed model on Site. ICV for both volumes and thicknesses. “Model” 1 in Mackey 2019. |
| Van Erp 2016 [12] | ROI ~ Diagnosis + Sex + Age + ICV + Site | Volumes | Bonferroni | “Model 1” in Van Erp 2016. |
| Van Erp 2018 [13] | ROI ~ Diagnosis + Sex + Age + Site | Thicknesses | False discovery rate | “Model A” in Van Erp 2018. |
| Schmaal 2016 [14] | ROI ~ Diagnosis + Sex + Age + ICV + Site | Volumes | Bonferroni | None. |
| Schmaal 2017 [15] | ROI ~ Diagnosis + Sex + Age + Site | Thicknesses | False discovery rate | None. |
| Groenewold 2023 [16] | ROI ~ Diagnosis + Sex + Age + $Age^2$ + Sex×Age + Sex×$Age^2$ + ICV + (1\|Site) | Volumes | Family-wise error rate | Mixed model on Site. |
| Present study | ROI ~ Diagnosis + Sex + Age + [ICV] + Site | Volumes and thicknesses | Bonferroni | ICV only for volumes. |

**Supplementary Table S10. Site distribution of Mental Disorders (any) patients and Non-Psychiatric Controls.** Sites with five or fewer patients are reported as "≤5" to reduce the risk of patient identification.

| Site | | Population A | | Population B | | Population C | |
|---|---|---|---|---|---|---|---|
| **Performing Hospital** | **Manufacturer Model Name** | **Mental Disorders (any)** | **Non-Psychiatric Controls** | **Mental Disorders (any)** | **Non-Psychiatric Controls** | **Mental Disorders (any)** | **Non-Psychiatric Controls** |
| AHH, AMAGER OG HVIDOVRE HOSPITAL | Avanto | 53 | 9 | 53 | ≤5 | 8 | ≤5 |
| | Prisma_fit | 52 | 6 | 52 | ≤5 | 16 | ≤5 |
| | Verio | 70 | 7 | 70 | ≤5 | 10 | ≤5 |
| BFH, BISPEBJERG OG FREDERIKSBERG HOSPITALER | Ingenia | 330 | 338 | 330 | 221 | 117 | 221 |
| | Ingenia Ambition X | 31 | 10 | 31 | ≤5 | 8 | ≤5 |
| | Verio | 199 | 24 | 199 | 103 | 85 | 103 |
| BOH, BORNHOLMS HOSPITAL | Aera | 45 | 30 | 45 | 20 | 8 | 20 |
| HGH, HERLEV OG GENTOFTE HOSPITAL | Achieva | 293 | 212 | 293 | 124 | 93 | 124 |
| | Ingenia | 108 | 92 | 108 | 47 | 26 | 47 |
| | Ingenia Elition X | 66 | 50 | 66 | 28 | 22 | 28 |
| | Panorama HFO | 10 | ≤5 | - | - | - | - |
| | SIGNA Artist | 11 | ≤5 | 11 | ≤5 | ≤5 | ≤5 |
| | Signa HDxt | 40 | 15 | 40 | 13 | 10 | 13 |
| NOH, HOSPITALERNE I NORDSJÆLLAND | Achieva dStream | 256 | 251 | 256 | 146 | 69 | 146 |
| | Ingenia | 262 | 237 | 262 | 120 | 80 | 120 |
| RH, RIGSHOSPITALET | Achieva dStream | 58 | 53 | 58 | 42 | 18 | 42 |
| | Avanto | 382 | 204 | 382 | 152 | 96 | 152 |
| | Avanto_fit | 88 | 54 | 88 | 21 | 15 | 21 |
| | ClinApps | 8 | 8 | 8 | ≤5 | ≤5 | ≤5 |
| | DISCOVERY MR450 | 23 | 12 | 23 | 7 | ≤5 | 7 |
| | Optima MR450w | 145 | 44 | 145 | 31 | 24 | 31 |
| | Prisma_fit | 42 | 25 | 42 | 18 | 7 | 18 |
| | Signa HDxt | ≤5 | 6 | ≤5 | ≤5 | ≤5 | ≤5 |
| | Verio | 48 | 26 | 48 | 17 | ≤5 | 17 |
| SJ, REGION SJÆLLANDS SYGEHUSVÆSEN | Aera | ≤5 | 9 | ≤5 | ≤5 | ≤5 | ≤5 |
| | Avanto_fit | 8 | 31 | 8 | 9 | ≤5 | 9 |
| Unknown | Achieva dStream | ≤5 | 23 | ≤5 | 16 | ≤5 | 16 |
| | Aera | ≤5 | 16 | ≤5 | 11 | ≤5 | 11 |
| | Avanto | 15 | 29 | 15 | 26 | 11 | 26 |
| | Avanto_fit | 12 | 34 | 12 | 22 | ≤5 | 22 |
| | Ingenia | ≤5 | 15 | ≤5 | 15 | ≤5 | 15 |
| | Prisma_fit | 31 | 129 | 31 | 106 | 18 | 106 |
| | Signa HDxt | ≤5 | ≤5 | ≤5 | ≤5 | - | - |
| | Skyra | ≤5 | ≤5 | ≤5 | ≤5 | - | - |
| | Verio | 24 | 27 | 24 | 15 | ≤5 | 15 |
| | Unknown | 43 | 25 | 43 | 13 | 12 | 13 |
| **TOTAL** | | **2774** | **2062** | **2764** | **1382** | **785** | **1376** |

**Supplementary Table S11. Site distribution of Mental Disorders without Dementia and Non-Psychiatric Controls.** Sites with five or fewer patients are reported as "≤5" to reduce the risk of patient identification.

| Site | | Population A | | Population B | | Population C | |
|---|---|---|---|---|---|---|---|
| **Performing Hospital** | **Manufacturer Model Name** | **Mental Disorders without Dementia** | **Non-Psychiatric Controls** | **Mental Disorders without Dementia** | **Non-Psychiatric Controls** | **Mental Disorders without Dementia** | **Non-Psychiatric Controls** |
| AHH, AMAGER OG HVIDOVRE HOSPITAL | Avanto | 52 | 9 | 52 | ≤5 | 8 | ≤5 |
| | Prisma_fit | 51 | 6 | 51 | ≤5 | 16 | ≤5 |
| | Verio | 67 | 7 | 67 | ≤5 | 9 | ≤5 |
| BFH, BISPEBJERG OG FREDERIKSBERG HOSPITALER | Ingenia | 317 | 338 | 317 | 221 | 116 | 221 |
| | Ingenia Ambition X | 28 | 10 | 28 | ≤5 | 6 | ≤5 |
| | Verio | 185 | 24 | 185 | 103 | 83 | 103 |
| BOH, BORNHOLMS HOSPITAL | Aera | 39 | 30 | 39 | 20 | 8 | 20 |
| HGH, HERLEV OG GENTOFTE HOSPITAL | Achieva | 274 | 212 | 274 | 124 | 87 | 124 |
| | Ingenia | 101 | 92 | 101 | 47 | 25 | 47 |
| | Ingenia Elition X | 66 | 50 | 66 | 28 | 22 | 28 |
| | Panorama HFO | 10 | ≤5 | - | - | - | - |
| | SIGNA Artist | 11 | ≤5 | 11 | ≤5 | ≤5 | ≤5 |
| | Signa HDxt | 38 | 15 | 38 | 13 | 9 | 13 |
| NOH, HOSPITALERNE I NORDSJÆLLAND | Achieva dStream | 231 | 251 | 231 | 146 | 65 | 146 |
| | Ingenia | 242 | 237 | 242 | 120 | 74 | 120 |
| RH, RIGSHOSPITALET | Achieva dStream | 58 | 53 | 58 | 42 | 18 | 42 |
| | Avanto | 371 | 204 | 371 | 152 | 95 | 152 |
| | Avanto_fit | 87 | 54 | 87 | 21 | 15 | 21 |
| | ClinApps | 6 | 8 | 6 | ≤5 | ≤5 | ≤5 |
| | DISCOVERY MR450 | 22 | 12 | 22 | 7 | ≤5 | 7 |
| | Optima MR450w | 128 | 44 | 128 | 31 | 20 | 31 |
| | Prisma_fit | 42 | 25 | 42 | 18 | 7 | 18 |
| | Signa HDxt | ≤5 | 6 | ≤5 | ≤5 | ≤5 | ≤5 |
| | Verio | 47 | 26 | 47 | 17 | ≤5 | 17 |
| SJ, REGION SJÆLLANDS SYGEHUSVÆSEN | Aera | ≤5 | 9 | ≤5 | ≤5 | ≤5 | ≤5 |
| | Avanto_fit | 8 | 31 | 8 | 9 | ≤5 | 9 |
| Unknown | Achieva dStream | ≤5 | 23 | ≤5 | 16 | ≤5 | 16 |
| | Aera | ≤5 | 16 | ≤5 | 11 | ≤5 | 11 |
| | Avanto | 15 | 29 | 15 | 26 | 11 | 26 |
| | Avanto_fit | 11 | 34 | 11 | 22 | ≤5 | 22 |
| | Ingenia | ≤5 | 15 | ≤5 | 15 | ≤5 | 15 |
| | Prisma_fit | 31 | 129 | 31 | 106 | 18 | 106 |
| | Signa HDxt | ≤5 | ≤5 | ≤5 | ≤5 | - | - |
| | Skyra | ≤5 | ≤5 | ≤5 | ≤5 | - | - |
| | Verio | 24 | 27 | 24 | 15 | ≤5 | 15 |
| | Unknown | 23 | 25 | 23 | 13 | 8 | 13 |
| **TOTAL** | | **2606** | **2062** | **2596** | **1382** | **751** | **1376** |

**Supplementary Table S12. Site distribution of Dementia patients and Non-Psychiatric Controls.** Sites with five or fewer patients are reported as "≤5" to reduce the risk of patient identification.

| Site | | Population A | | Population B | | Population C | |
|---|---|---|---|---|---|---|---|
| **Performing Hospital** | **Manufacturer Model Name** | **Dementia** | **Non-Psychiatric Controls** | **Dementia** | **Non-Psychiatric Controls** | **Dementia** | **Non-Psychiatric Controls** |
| AHH, AMAGER OG HVIDOVRE HOSPITAL | Avanto | ≤5 | 9 | ≤5 | ≤5 | - | - |
| | Prisma_fit | ≤5 | 6 | ≤5 | ≤5 | - | - |
| | Verio | ≤5 | 7 | ≤5 | ≤5 | ≤5 | ≤5 |
| BFH, BISPEBJERG OG FREDERIKSBERG HOSPITALER | Ingenia | 15 | 338 | 15 | 221 | ≤5 | 221 |
| | Ingenia Ambition X | ≤5 | 10 | ≤5 | ≤5 | ≤5 | ≤5 |
| | Verio | 15 | 24 | 15 | 103 | ≤5 | 103 |
| BOH, BORNHOLMS HOSPITAL | Aera | 13 | 30 | 13 | 20 | ≤5 | 20 |
| HGH, HERLEV OG GENTOFTE HOSPITAL | Achieva | 31 | 212 | 31 | 124 | 6 | 124 |
| | Ingenia | 7 | 92 | 7 | 47 | ≤5 | 47 |
| | Ingenia Elition X | ≤5 | 50 | ≤5 | 28 | - | - |
| | Panorama HFO | - | - | - | - | - | - |
| | SIGNA Artist | - | - | - | - | - | - |
| | Signa HDxt | ≤5 | 15 | ≤5 | 13 | ≤5 | 13 |
| NOH, HOSPITALERNE I NORDSJÆLLAND | Achieva dStream | 28 | 251 | 28 | 146 | 6 | 146 |
| | Ingenia | 28 | 237 | 28 | 120 | 8 | 120 |
| RH, RIGSHOSPITALET | Achieva dStream | ≤5 | 53 | ≤5 | 42 | - | - |
| | Avanto | 29 | 204 | 29 | 152 | ≤5 | 152 |
| | Avanto_fit | ≤5 | 54 | ≤5 | 21 | - | - |
| | ClinApps | ≤5 | 8 | ≤5 | ≤5 | - | - |
| | DISCOVERY MR450 | ≤5 | 12 | ≤5 | 7 | - | - |
| | Optima MR450w | 26 | 44 | 26 | 31 | 7 | 31 |
| | Prisma_fit | ≤5 | 25 | ≤5 | 18 | ≤5 | 18 |
| | Signa HDxt | ≤5 | 6 | ≤5 | ≤5 | - | - |
| | Verio | ≤5 | 26 | ≤5 | 17 | - | - |
| SJ, REGION SJÆLLANDS SYGEHUSVÆSEN | Aera | - | - | - | - | - | - |
| | Avanto_fit | ≤5 | 31 | ≤5 | 9 | ≤5 | 9 |
| Unknown | Achieva dStream | - | - | - | - | - | - |
| | Aera | - | - | - | - | - | - |
| | Avanto | - | - | - | - | - | - |
| | Avanto_fit | ≤5 | 34 | ≤5 | 22 | ≤5 | 22 |
| | Ingenia | ≤5 | 15 | ≤5 | 15 | - | - |
| | Prisma_fit | - | - | - | - | - | - |
| | Signa HDxt | - | - | - | - | - | - |
| | Skyra | - | - | - | - | - | - |
| | Verio | - | - | - | - | - | - |
| | Unknown | 41 | 25 | 41 | 13 | 13 | 13 |
| **TOTAL** | | **262** | **1818** | **262** | **1194** | **60** | **1048** |

**Supplementary Table S13. Site distribution of Substance Use Disorder patients and Non-Psychiatric Controls.** Sites with five or fewer patients are reported as "≤5" to reduce the risk of patient identification.

| Site | | Population A | | Population B | | Population C | |
|---|---|---|---|---|---|---|---|
| **Performing Hospital** | **Manufacturer Model Name** | **Substance Use Disorder** | **Non-Psychiatric Controls** | **Substance Use Disorder** | **Non-Psychiatric Controls** | **Substance Use Disorder** | **Non-Psychiatric Controls** |
| AHH, AMAGER OG HVIDOVRE HOSPITAL | Avanto | 7 | 9 | 7 | ≤5 | ≤5 | ≤5 |
| | Prisma_fit | 11 | 6 | 11 | ≤5 | ≤5 | ≤5 |
| | Verio | ≤5 | 7 | ≤5 | ≤5 | ≤5 | ≤5 |
| BFH, BISPEBJERG OG FREDERIKSBERG HOSPITALER | Ingenia | 76 | 338 | 76 | 221 | 17 | 221 |
| | Ingenia Ambition X | ≤5 | 10 | ≤5 | ≤5 | ≤5 | ≤5 |
| | Verio | 41 | 24 | 41 | 103 | 16 | 103 |
| BOH, BORNHOLMS HOSPITAL | Aera | ≤5 | 30 | ≤5 | 20 | - | - |
| HGH, HERLEV OG GENTOFTE HOSPITAL | Achieva | 68 | 212 | 68 | 124 | 17 | 124 |
| | Ingenia | 28 | 92 | 28 | 47 | ≤5 | 47 |
| | Ingenia Elition X | 20 | 50 | 20 | 28 | ≤5 | 28 |
| | Panorama HFO | ≤5 | ≤5 | - | - | - | - |
| | SIGNA Artist | ≤5 | ≤5 | ≤5 | ≤5 | ≤5 | ≤5 |
| | Signa HDxt | 17 | 15 | 17 | 13 | ≤5 | 13 |
| NOH, HOSPITALERNE I NORDSJÆLLAND | Achieva dStream | 48 | 251 | 48 | 146 | 11 | 146 |
| | Ingenia | 67 | 237 | 67 | 120 | 17 | 120 |
| RH, RIGSHOSPITALET | Achieva dStream | 10 | 53 | 10 | 42 | ≤5 | 42 |
| | Avanto | 62 | 204 | 62 | 152 | ≤5 | 152 |
| | Avanto_fit | 15 | 54 | 15 | 21 | ≤5 | 21 |
| | ClinApps | ≤5 | 8 | ≤5 | ≤5 | - | - |
| | DISCOVERY MR450 | ≤5 | 12 | ≤5 | 7 | - | - |
| | Optima MR450w | 29 | 44 | 29 | 31 | ≤5 | 31 |
| | Prisma_fit | 8 | 25 | 8 | 18 | - | - |
| | Signa HDxt | ≤5 | 6 | ≤5 | ≤5 | - | - |
| | Verio | 11 | 26 | 11 | 17 | - | - |
| SJ, REGION SJÆLLANDS SYGEHUSVÆSEN | Aera | ≤5 | 9 | ≤5 | ≤5 | ≤5 | ≤5 |
| | Avanto_fit | ≤5 | 31 | ≤5 | 9 | - | - |
| Unknown | Achieva dStream | - | - | - | - | - | - |
| | Aera | ≤5 | 16 | ≤5 | 11 | - | - |
| | Avanto | - | - | - | - | - | - |
| | Avanto_fit | ≤5 | 34 | ≤5 | 22 | ≤5 | 22 |
| | Ingenia | ≤5 | 15 | ≤5 | 15 | - | - |
| | Prisma_fit | 11 | 129 | 11 | 106 | 7 | 106 |
| | Signa HDxt | ≤5 | ≤5 | ≤5 | ≤5 | - | - |
| | Skyra | - | - | - | - | - | - |
| | Verio | ≤5 | 27 | ≤5 | 15 | - | - |
| | Unknown | ≤5 | 25 | ≤5 | 13 | - | - |
| **TOTAL** | | **574** | **2005** | **569** | **1335** | **119** | **1201** |

**Supplementary Table S14. Site distribution of Schizophrenia Spectrum Disorder patients and Non-Psychiatric Controls.** Sites with five or fewer patients are reported as "≤5" to reduce the risk of patient identification.

| Site | | Population A | | Population B | | Population C | |
|---|---|---|---|---|---|---|---|
| **Performing Hospital** | **Manufacturer Model Name** | **Schizo-phrenia Spectrum Disorder** | **Non-Psychiatric Controls** | **Schizo-phrenia Spectrum Disorder** | **Non-Psychiatric Controls** | **Schizo-phrenia Spectrum Disorder** | **Non-Psychiatric Controls** |
| AHH, AMAGER OG HVIDOVRE HOSPITAL | Avanto | 11 | 9 | 11 | ≤5 | ≤5 | ≤5 |
| | Prisma_fit | 16 | 6 | 16 | ≤5 | 10 | ≤5 |
| | Verio | 16 | 7 | 16 | ≤5 | ≤5 | ≤5 |
| BFH, BISPEBJERG OG FREDERIKSBERG HOSPITALER | Ingenia | 56 | 338 | 56 | 221 | 31 | 221 |
| | Ingenia Ambition X | ≤5 | 10 | ≤5 | ≤5 | ≤5 | ≤5 |
| | Verio | 48 | 24 | 48 | 103 | 33 | 103 |
| BOH, BORNHOLMS HOSPITAL | Aera | ≤5 | 30 | ≤5 | 20 | ≤5 | 20 |
| HGH, HERLEV OG GENTOFTE HOSPITAL | Achieva | 49 | 212 | 49 | 124 | 25 | 124 |
| | Ingenia | 17 | 92 | 17 | 47 | 9 | 47 |
| | Ingenia Elition X | 10 | 50 | 10 | 28 | 6 | 28 |
| | Panorama HFO | - | - | - | - | - | - |
| | SIGNA Artist | ≤5 | ≤5 | ≤5 | ≤5 | ≤5 | ≤5 |
| | Signa HDxt | ≤5 | 15 | ≤5 | 13 | ≤5 | 13 |
| NOH, HOSPITALERNE I NORDSJÆLLAND | Achieva dStream | 24 | 251 | 24 | 146 | 11 | 146 |
| | Ingenia | 32 | 237 | 32 | 120 | 12 | 120 |
| RH, RIGSHOSPITALET | Achieva dStream | 19 | 53 | 19 | 42 | 10 | 42 |
| | Avanto | 66 | 204 | 66 | 152 | 36 | 152 |
| | Avanto_fit | 12 | 54 | 12 | 21 | ≤5 | 21 |
| | ClinApps | - | - | - | - | - | - |
| | DISCOVERY MR450 | ≤5 | 12 | ≤5 | 7 | - | - |
| | Optima MR450w | 19 | 44 | 19 | 31 | 6 | 31 |
| | Prisma_fit | 6 | 25 | 6 | 18 | ≤5 | 18 |
| | Signa HDxt | - | - | - | - | - | - |
| | Verio | 8 | 26 | 8 | 17 | ≤5 | 17 |
| SJ, REGION SJÆLLANDS SYGEHUSVÆSEN | Aera | ≤5 | 9 | ≤5 | ≤5 | - | - |
| | Avanto_fit | ≤5 | 31 | ≤5 | 9 | - | - |
| Unknown | Achieva dStream | - | - | - | - | - | - |
| | Aera | - | - | - | - | - | - |
| | Avanto | ≤5 | 29 | ≤5 | 26 | ≤5 | 26 |
| | Avanto_fit | ≤5 | 34 | ≤5 | 22 | - | - |
| | Ingenia | - | - | - | - | - | - |
| | Prisma_fit | ≤5 | 129 | ≤5 | 106 | ≤5 | 106 |
| | Signa HDxt | - | - | - | - | - | - |
| | Skyra | - | - | - | - | - | - |
| | Verio | ≤5 | 27 | ≤5 | 15 | - | - |
| | Unknown | ≤5 | 25 | ≤5 | 13 | ≤5 | 13 |
| **TOTAL** | | **435** | **1987** | **435** | **1326** | **214** | **1269** |

**Supplementary Table S15. Site distribution of Depression patients and Non-Psychiatric Controls.** Sites with five or fewer patients are reported as "≤5" to reduce the risk of patient identification.

| Site | | Population A | | Population B | | Population C | |
|---|---|---|---|---|---|---|---|
| **Performing Hospital** | **Manufacturer Model Name** | **Depression** | **Non-Psychiatric Controls** | **Depression** | **Non-Psychiatric Controls** | **Depression** | **Non-Psychiatric Controls** |
| AHH, AMAGER OG HVIDOVRE HOSPITAL | Avanto | 13 | 9 | 13 | ≤5 | - | - |
| | Prisma_fit | 9 | 6 | 9 | ≤5 | ≤5 | ≤5 |
| | Verio | 12 | 7 | 12 | ≤5 | ≤5 | ≤5 |
| BFH, BISPEBJERG OG FREDERIKSBERG HOSPITALER | Ingenia | 70 | 338 | 70 | 221 | 25 | 221 |
| | Ingenia Ambition X | 9 | 10 | 9 | ≤5 | ≤5 | ≤5 |
| | Verio | 50 | 24 | 50 | 103 | 18 | 103 |
| BOH, BORNHOLMS HOSPITAL | Aera | 8 | 30 | 8 | 20 | ≤5 | 20 |
| HGH, HERLEV OG GENTOFTE HOSPITAL | Achieva | 77 | 212 | 77 | 124 | 24 | 124 |
| | Ingenia | 27 | 92 | 27 | 47 | 7 | 47 |
| | Ingenia Elition X | 13 | 50 | 13 | 28 | ≤5 | 28 |
| | Panorama HFO | ≤5 | ≤5 | - | - | - | - |
| | SIGNA Artist | ≤5 | ≤5 | ≤5 | ≤5 | - | - |
| | Signa HDxt | ≤5 | 15 | ≤5 | 13 | - | - |
| NOH, HOSPITALERNE I NORDSJÆLLAND | Achieva dStream | 72 | 251 | 72 | 146 | 19 | 146 |
| | Ingenia | 59 | 237 | 59 | 120 | 18 | 120 |
| RH, RIGSHOSPITALET | Achieva dStream | 15 | 53 | 15 | 42 | ≤5 | 42 |
| | Avanto | 103 | 204 | 103 | 152 | 24 | 152 |
| | Avanto_fit | 20 | 54 | 20 | 21 | ≤5 | 21 |
| | ClinApps | ≤5 | 8 | ≤5 | ≤5 | ≤5 | ≤5 |
| | DISCOVERY MR450 | 12 | 12 | 12 | 7 | ≤5 | 7 |
| | Optima MR450w | 36 | 44 | 36 | 31 | ≤5 | 31 |
| | Prisma_fit | 9 | 25 | 9 | 18 | ≤5 | 18 |
| | Signa HDxt | ≤5 | 6 | ≤5 | ≤5 | ≤5 | ≤5 |
| | Verio | 16 | 26 | 16 | 17 | ≤5 | ≤5 |
| SJ, REGION SJÆLLANDS SYGEHUSVÆSEN | Aera | - | - | - | - | - | - |
| | Avanto_fit | ≤5 | 31 | ≤5 | 9 | ≤5 | 9 |
| Unknown | Achieva dStream | ≤5 | 23 | ≤5 | 16 | ≤5 | 16 |
| | Aera | ≤5 | 16 | ≤5 | 11 | ≤5 | 11 |
| | Avanto | ≤5 | 29 | ≤5 | 26 | ≤5 | 26 |
| | Avanto_fit | ≤5 | 34 | ≤5 | 22 | ≤5 | 22 |
| | Ingenia | ≤5 | 15 | ≤5 | 15 | ≤5 | 15 |
| | Prisma_fit | 10 | 129 | 10 | 106 | ≤5 | 106 |
| | Signa HDxt | - | - | - | - | - | - |
| | Skyra | - | - | - | - | - | - |
| | Verio | ≤5 | 27 | ≤5 | 15 | - | - |
| | Unknown | ≤5 | 25 | ≤5 | 13 | ≤5 | 13 |
| **TOTAL** | | **676** | **2047** | **674** | **1372** | **179** | **1335** |

**Supplementary Table S16. Site distribution of Anxiety Disorder patients and Non-Psychiatric Controls.** Sites with five or fewer patients are reported as "≤5" to reduce the risk of patient identification.

| **Site** | | **Population A** | | **Population B** | | **Population C** | |
|---|---|---|---|---|---|---|---|
| **Performing Hospital** | **Manufacturer Model Name** | **Anxiety Disorder** | **Non-Psychiatric Controls** | **Anxiety Disorder** | **Non-Psychiatric Controls** | **Anxiety Disorder** | **Non-Psychiatric Controls** |
| AHH, AMAGER OG HVIDOVRE HOSPITAL | Avanto | 12 | 9 | 12 | ≤5 | - | - |
| | Prisma_fit | 7 | 6 | 7 | ≤5 | ≤5 | ≤5 |
| | Verio | 17 | 7 | 17 | ≤5 | - | - |
| BFH, BISPEBJERG OG FREDERIKSBERG HOSPITALER | Ingenia | 67 | 338 | 67 | 221 | 27 | 221 |
| | Ingenia Ambition X | ≤5 | 10 | ≤5 | ≤5 | ≤5 | ≤5 |
| | Verio | 30 | 24 | 30 | 103 | 11 | 103 |
| BOH, BORNHOLMS HOSPITAL | Aera | 12 | 30 | 12 | 20 | ≤5 | 20 |
| HGH, HERLEV OG GENTOFTE HOSPITAL | Achieva | 41 | 212 | 41 | 124 | 13 | 124 |
| | Ingenia | 19 | 92 | 19 | 47 | ≤5 | 47 |
| | Ingenia Elition X | 11 | 50 | 11 | 28 | ≤5 | 28 |
| | Panorama HFO | - | - | - | - | - | - |
| | SIGNA Artist | ≤5 | ≤5 | ≤5 | ≤5 | - | - |
| | Signa HDxt | 8 | 15 | 8 | 13 | ≤5 | 13 |
| NOH, HOSPITALERNE I NORDSJÆLLAND | Achieva dStream | 53 | 251 | 53 | 146 | 17 | 146 |
| | Ingenia | 58 | 237 | 58 | 120 | 22 | 120 |
| RH, RIGSHOSPITALET | Achieva dStream | 7 | 53 | 7 | 42 | ≤5 | 42 |
| | Avanto | 80 | 204 | 80 | 152 | 20 | 152 |
| | Avanto_fit | 17 | 54 | 17 | 21 | ≤5 | 21 |
| | ClinApps | ≤5 | 8 | ≤5 | ≤5 | ≤5 | ≤5 |
| | DISCOVERY MR450 | ≤5 | 12 | ≤5 | 7 | ≤5 | 7 |
| | Optima MR450w | 22 | 44 | 22 | 31 | ≤5 | 31 |
| | Prisma_fit | 8 | 25 | 8 | 18 | ≤5 | 18 |
| | Signa HDxt | ≤5 | 6 | ≤5 | ≤5 | - | - |
| | Verio | 7 | 26 | 7 | 17 | - | - |
| SJ, REGION SJÆLLANDS SYGEHUSVÆSEN | Aera | ≤5 | 9 | ≤5 | 9 | - | - |
| | Avanto_fit | ≤5 | 31 | ≤5 | ≤5 | ≤5 | 9 |
| Unknown | Achieva dStream | ≤5 | 23 | ≤5 | 16 | ≤5 | 16 |
| | Aera | - | - | - | - | - | - |
| | Avanto | 7 | 29 | 7 | 26 | 6 | 26 |
| | Avanto_fit | ≤5 | 34 | ≤5 | 22 | - | - |
| | Ingenia | - | - | - | - | - | - |
| | Prisma_fit | 8 | 129 | 8 | 106 | ≤5 | 106 |
| | Signa HDxt | - | - | - | - | - | - |
| | Skyra | - | - | - | - | - | - |
| | Verio | 9 | 27 | 9 | 15 | ≤5 | 15 |
| | Unknown | ≤5 | 25 | ≤5 | 13 | ≤5 | 13 |
| **TOTAL** | | **524** | **2024** | **524** | **1350** | **155** | **1289** |

**Supplementary Table S17: Comparison of Mental Disorders and Dementia patients for previously reported ROI differences for Population A.**

| Population A | Cohen's d [95% CI] | p-value |
|---|---|---|
| **Non-Psychiatric Controls (N=2062) vs. Mental Disorders (N=2774)** | | |
| Lateral Ventricles | 0.334 [0.278, 0.391] | 3.40e-30 * |
| Thalamus | -0.364 [-0.421, -0.307] | 2.23e-35 * |
| Amygdala | -0.268 [-0.325, -0.211] | 4.48e-20 * |
| Hippocampus | -0.167 [-0.224, -0.111] | 9.51e-09 * |
| Cortical Thickness | -0.010 [-0.066, 0.047] | 7.36e-01 |
| Pars Opercularis | -0.027 [-0.083, 0.030] | 3.58e-01 |
| Medial Orbitofrontal | -0.051 [-0.108, -0.005] | 7.87e-02 |
| Rostral Anterior Cingulate | -0.060 [-0.117, -0.004] | 3.85e-02 |
| Superior Temporal | 0.019 [-0.037, 0.075] | 5.16e-01 |
| Insula | -0.127 [-0.184, -0.071] | 1.25e-05 * |
| **Non-Psychiatric Controls (N=1818) vs. Dementia (N=262)** | | |
| Lateral Ventricles | 0.886 [0.795, 0.976] | 2.56e-39 * |
| Hippocampus | -0.603 [-0.691, -0.515] | 1.65e-19 * |
| Cortical Thickness | -0.302 [-0.388, -0.215] | 5.33e-06 * |
| Entorhinal | -0.487 [-0.574, -0.399] | 2.57e-13 * |
| **Non-Psychiatric Controls (N=2062) vs. Mental Disorders without Dementia (N=2606)** | | |
| Lateral Ventricles | 0.303 [0.245, 0.361] | 1.54e-24 * |
| Thalamus | -0.353 [-0.411, -0.295] | 1.32e-32 * |
| Amygdala | -0.248 [-0.305, -0.190] | 5.46e-17 * |
| Hippocampus | -0.149 [-0.206, -0.092] | 4.46e-07 * |
| Cortical Thickness | -0.001 [-0.058, 0.056] | 9.72e-01 |
| Pars Opercularis | -0.023 [-0.080, 0.035] | 4.40e-01 |
| Medial Orbitofrontal | -0.045 [-0.102, 0.013] | 1.29e-01 |
| Rostral Anterior Cingulate | -0.056 [-0.114, -0.001] | 5.56e-02 |
| Superior Temporal | 0.027 [-0.031, 0.084] | 3.65e-01 |
| Insula | -0.118 [-0.175, -0.060] | 6.76e-05 * |

**Supplementary Table S18: Comparison of Mental Disorders and Dementia patients for previously reported ROI differences for Population B.**

| Population B | Cohen's d [95% CI] | p-value |
|---|---|---|
| **Non-Psychiatric Controls (N=1382) vs. Mental Disorders (N=2764)** | | |
| Lateral Ventricles | 0.376 [0.314, 0.437] | 1.18e-29 * |
| Thalamus | -0.394 [-0.456, -0.333] | 1.74e-32 * |
| Amygdala | -0.283 [-0.344, -0.222] | 1.16e-17 * |
| Hippocampus | -0.181 [-0.242, -0.120] | 4.25e-08 * |
| Cortical Thickness | -0.044 [-0.105, 0.017] | 1.78e-01 |
| Pars Opercularis | -0.061 [-0.122, 0.000] | 6.53e-02 |
| Medial Orbitofrontal | -0.062 [-0.123, -0.001] | 5.88e-02 |
| Rostral Anterior Cingulate | -0.093 [-0.154, -0.032] | 4.90e-03 * |
| Superior Temporal | -0.031 [-0.092, 0.030] | 3.44e-01 |
| Insula | -0.129 [-0.190, -0.068] | 9.66e-05 * |
| **Non-Psychiatric Controls (N=1194) vs. Dementia (N=262)** | | |
| Lateral Ventricles | 1.037 [0.927, 1.146] | 1.84e-48 * |
| Hippocampus | -0.651 [-0.757, -0.546] | 5.80e-21 * |
| Cortical Thickness | -0.333 [-0.437, -0.230] | 1.16e-06 * |
| Entorhinal | -0.552 [-0.657, -0.448] | 1.21e-15 * |
| **Non-Psychiatric Controls (N=1382) vs. Mental Disorders without Dementia (N=2596)** | | |
| Lateral Ventricles | 0.351 [0.289, 0.414] | 1.12e-25 * |
| Thalamus | -0.384 [-0.447, -0.321] | 2.72e-30 * |
| Amygdala | -0.267 [-0.329, -0.204] | 1.51e-15 * |
| Hippocampus | -0.165 [-0.227, -0.103] | 7.53e-07 * |
| Cortical Thickness | -0.037 [-0.099, 0.026] | 2.73e-01 |
| Pars Opercularis | -0.058 [-0.120, -0.005] | 8.41e-02 |
| Medial Orbitofrontal | -0.056 [-0.118, -0.007] | 9.48e-02 |
| Rostral Anterior Cingulate | -0.087 [-0.149, -0.025] | 9.07e-03 |
| Superior Temporal | -0.024 [-0.086, 0.038] | 4.75e-01 |
| Insula | -0.120 [-0.182, -0.057] | 3.36e-04 * |

**Supplementary Table S19: Comparison of Mental Disorders and Dementia patients for previously reported ROI differences for Population C.**

| Population C | Cohen's d [95% CI] | p-value |
|---|---|---|
| **Non-Psychiatric Controls (N=1376) vs. Mental Disorders (N=785)** | | |
| Lateral Ventricles | 0.272 [0.187, 0.357] | 1.46e-09 * |
| Thalamus | -0.298 [-0.383, -0.213] | 3.35e-11 * |
| Amygdala | -0.250 [-0.335, -0.165] | 2.54e-08 * |
| Hippocampus | -0.145 [-0.229, -0.060] | 1.25e-03 * |
| Cortical Thickness | -0.180 [-0.264, -0.095] | 5.98e-05 * |
| Pars Opercularis | -0.158 [-0.242, -0.073] | 4.37e-04 * |
| Medial Orbitofrontal | -0.132 [-0.216, -0.047] | 3.24e-03 * |
| Rostral Anterior Cingulate | -0.124 [-0.208, -0.039] | 5.65e-03 |
| Superior Temporal | -0.151 [-0.235, -0.066] | 7.62e-04 * |
| Insula | -0.177 [-0.262, -0.093] | 7.67e-05 * |
| **Non-Psychiatric Controls (N=1048) vs. Dementia (N=60)** | | |
| Lateral Ventricles | 0.757 [0.635, 0.879] | 1.53e-08 * |
| Hippocampus | -0.632 [-0.753, -0.512] | 2.15e-06 * |
| Cortical Thickness | -0.648 [-0.769, -0.527] | 1.21e-06 * |
| Entorhinal | -0.690 [-0.811, -0.569] | 2.41e-07 * |
| **Non-Psychiatric Controls (N=1376) vs. Mental Disorders without Dementia (N=751)** | | |
| Lateral Ventricles | 0.247 [0.162, 0.333] | 5.55e-08 * |
| Thalamus | -0.283 [-0.369, -0.198] | 5.15e-10 * |
| Amygdala | -0.217 [-0.302, -0.132] | 1.89e-06 * |
| Hippocampus | -0.125 [-0.210, -0.039] | 6.11e-03 |
| Cortical Thickness | -0.160 [-0.245, -0.074] | 4.46e-04 * |
| Pars Opercularis | -0.146 [-0.231, -0.060] | 1.35e-03 * |
| Medial Orbitofrontal | -0.122 [-0.207, -0.037] | 7.31e-03 |
| Rostral Anterior Cingulate | -0.113 [-0.198, -0.028] | 1.26e-02 |
| Superior Temporal | -0.141 [-0.226, -0.055] | 1.98e-03 * |
| Insula | -0.163 [-0.248, -0.078] | 3.32e-04 * |

**Supplementary Table S20: Comparison of findings for high- (≤1.5mm slice thickness) and low-resolution (>1.5mm slice thickness) scans for Population A.**

| Population A. Overlap: Non-Psychiatric Controls: 47.53% Mental Disorders: 40.62% | All Resolutions: Non-Psychiatric Controls (N=2062) Mental Disorders (N=2774) | | High Resolution: Non-Psychiatric Controls (N=1371) Mental Disorders (N=1655) | | Low Resolution: Non-Psychiatric Controls (N=1870) Mental Disorders (N=2246) | |
|---|---|---|---|---|---|---|
| | **Cohen's d [95% CI]** | **p-value** | **Cohen's d [95% CI]** | **p-value** | **Cohen's d [95% CI]** | **p-value** |
| Lateral Ventricles | 0.334 [0.278, 0.391] | 3.40e-30 * | 0.321 [0.250, 0.393] | 2.32e-18 * | 0.332 [0.270, 0.393] | 6.50e-26 * |
| Thalamus | -0.364 [-0.421, -0.307] | 2.23e-35 * | -0.365 [-0.436, -0.293] | 4.14e-23 * | -0.371 [-0.433, -0.310] | 6.45e-32 * |
| Amygdala | -0.268 [-0.325, -0.211] | 4.48e-20 * | -0.295 [-0.367, -0.224] | 8.95e-16 * | -0.278 [-0.339, -0.216] | 1.03e-18 * |
| Hippocampus | -0.167 [-0.224, -0.111] | 9.51e-09 * | -0.177 [-0.248, -0.105] | 1.40e-06 * | -0.185 [-0.246, -0.123] | 4.02e-09 * |
| Cortical Thickness | -0.010 [-0.066, 0.047] | 7.36e-01 | 0.051 [-0.020, 0.122] | 1.63e-01 | 0.074 [0.013, 0.135] | 1.77e-02 |
| Pars Opercularis | -0.027 [-0.083, 0.030] | 3.58e-01 | 0.004 [-0.075, 0.068] | 9.18e-01 | 0.024 [-0.037, 0.085] | 4.43e-01 |
| Medial Orbitofrontal | -0.051 [-0.108, -0.005] | 7.87e-02 | 0.122 [0.050, 0.193] | 8.77e-04 * | 0.043 [-0.018, 0.104] | 1.71e-01 |
| Rostral Anterior Cingulate | -0.060 [-0.117, -0.004] | 3.85e-02 | -0.003 [-0.074, 0.068] | 9.35e-01 | -0.041 [-0.102, 0.020] | 1.88e-01 |
| Superior Temporal | 0.019 [-0.037, 0.075] | 5.16e-01 | 0.053 [-0.018, 0.124] | 1.45e-01 | 0.085 [0.024, 0.146] | 6.72e-03 |
| Insula | -0.127 [-0.184, -0.071] | 1.25e-05 * | -0.092 [-0.163, -0.021] | 1.18e-02 | -0.074 [-0.135, -0.013] | 1.80e-02 |

**Supplementary Table S21: Comparison of findings for high- (≤1.5mm slice thickness) and low-resolution (>1.5mm slice thickness) scans for Population B.**

| Population B. Overlap: Non-Psychiatric Controls: 42.84% Mental Disorders: 40.38% | All Resolutions: Non-Psychiatric Controls (N=1382) Mental Disorders (N=2764) | | High Resolution: Non-Psychiatric Controls (N=822) Mental Disorders (N=1643) | | Low Resolution: Non-Psychiatric Controls (N=1152) Mental Disorders (N=2236) | |
|---|---|---|---|---|---|---|
| | Cohen's d [95% CI] | p-value | Cohen's d [95% CI] | p-value | Cohen's d [95% CI] | p-value |
| Lateral Ventricles | 0.376 [0.314, 0.437] | 1.18e-29 * | 0.383 [0.304, 0.463] | 5.70e-19 * | 0.372 [0.304, 0.440] | 2.73e-24 * |
| Thalamus | -0.394 [-0.456, -0.333] | 1.74e-32 * | -0.374 [-0.453, -0.294] | 3.98e-18 * | -0.403 [-0.472, -0.335] | 2.96e-28 * |
| Amygdala | -0.283 [-0.344, -0.222] | 1.16e-17 * | -0.286 [-0.366, -0.207] | 2.50e-11 * | -0.318 [-0.385, -0.250] | 3.18e-18 * |
| Hippocampus | -0.181 [-0.242, -0.120] | 4.25e-08 * | -0.183 [-0.263, -0.104] | 1.84e-05 * | -0.202 [-0.270, -0.134] | 2.76e-08 * |
| Cortical Thickness | -0.044 [-0.105, 0.017] | 1.78e-01 | 0.028 [-0.051, 0.107] | 5.13e-01 | 0.057 [-0.011, 0.124] | 1.17e-01 |
| Pars Opercularis | -0.061 [-0.122, 0.000] | 6.53e-02 | -0.035 [-0.114, 0.044] | 4.14e-01 | -0.003 [-0.071, 0.064] | 9.30e-01 |
| Medial Orbitofrontal | -0.062 [-0.123, -0.001] | 5.88e-02 | 0.125 [0.046, 0.204] | 3.42e-03 * | 0.043 [-0.024, 0.110] | 2.35e-01 |
| Rostral Anterior Cingulate | -0.093 [-0.154, -0.032] | 4.90e-03 * | -0.011 [-0.090, 0.068] | 7.92e-01 | -0.071 [-0.138, -0.004] | 5.01e-02 |
| Superior Temporal | -0.031 [-0.092, 0.030] | 3.44e-01 | 0.012 [-0.067, 0.091] | 7.86e-01 | 0.030 [-0.038, 0.097] | 4.11e-01 |
| Insula | -0.129 [-0.190, -0.068] | 9.66e-05 * | -0.083 [-0.162, -0.004] | 5.21e-02 | -0.080 [-0.147, -0.012] | 2.84e-02 |

**Supplementary Table S22: Comparison of findings for high- (≤1.5mm slice thickness) and low-resolution (>1.5mm slice thickness) scans for Population C.**

| Population C. Overlap: Non-Psychiatric Controls: 42.73% Mental Disorders: 38.98% | All Resolutions: Non-Psychiatric Controls (N=1376) Mental Disorders (N=785) | | High Resolution: Non-Psychiatric Controls (N=817) Mental Disorders (N=482) | | Low Resolution: Non-Psychiatric Controls (N=1147) Mental Disorders (N=609) | |
|---|---|---|---|---|---|---|
| | Cohen's d [95% CI] | p-value | Cohen's d [95% CI] | p-value | Cohen's d [95% CI] | p-value |
| Lateral Ventricles | 0.272 [0.187, 0.357] | 1.46e-09 * | 0.319 [0.210, 0.429] | 3.30e-08 * | 0.245 [0.151, 0.339] | 1.13e-06 * |
| Thalamus | -0.298 [-0.383, -0.213] | 3.35e-11 * | -0.281 [-0.391, -0.172] | 1.08e-06 * | -0.298 [-0.392, -0.204] | 3.37e-09 * |
| Amygdala | -0.250 [-0.335, -0.165] | 2.54e-08 * | -0.283 [-0.392, -0.173] | 9.66e-07 * | -0.271 [-0.365, -0.177] | 7.40e-08 * |
| Hippocampus | -0.145 [-0.229, -0.060] | 1.25e-03 * | -0.144 [-0.253, -0.035] | 1.22e-02 | -0.180 [-0.274, -0.086] | 3.40e-04 * |
| Cortical Thickness | -0.180 [-0.264, -0.095] | 5.98e-05 * | -0.136 [-0.245, -0.027] | 1.81e-02 | -0.082 [-0.176, 0.012] | 1.02e-01 |
| Pars Opercularis | -0.158 [-0.242, -0.073] | 4.37e-04 * | -0.129 [-0.238, -0.020] | 2.49e-02 | -0.092 [-0.186, 0.002] | 6.66e-02 |
| Medial Orbitofrontal | -0.132 [-0.216, -0.047] | 3.24e-03 * | 0.021 [-0.087, 0.130] | 7.10e-01 | -0.020 [-0.114, 0.074] | 6.91e-01 |
| Rostral Anterior Cingulate | -0.124 [-0.208, -0.039] | 5.65e-03 | -0.102 [-0.211, 0.007] | 7.65e-02 | -0.073 [-0.167, 0.021] | 1.46e-01 |
| Superior Temporal | -0.151 [-0.235, -0.066] | 7.62e-04 * | -0.123 [-0.232, -0.014] | 3.22e-02 | -0.086 [-0.180, 0.007] | 8.52e-02 |
| Insula | -0.177 [-0.262, -0.093] | 7.67e-05 * | -0.148 [-0.257, -0.039] | 1.03e-02 | -0.093 [-0.187, 0.000] | 6.32e-02 |

### Supplementary Table S23: Comparisons of all ROIs for Population A.

| Population A | Cohen's d [95% CI] | p-value |
|---|---|---|
| Non-Psychiatric Controls (N=2062) vs. Mental Disorders (N=2774) | | |
| CSF | 0.110 [0.054, 0.166] | 1.55e-04 * |
| Brain Stem | -0.188 [-0.244, -0.131] | 1.22e-10 * |
| 4th Ventricle | 0.124 [0.068, 0.181] | 2.01e-05 * |
| 3rd Ventricle | 0.352 [0.296, 0.409] | 2.59e-33 * |
| Ventral DC | -0.290 [-0.347, -0.233] | 3.37e-23 * |
| Nucleus Accumbens | -0.224 [-0.281, -0.168] | 1.43e-14 * |
| Amygdala | -0.268 [-0.325, -0.211] | 4.48e-20 * |
| Hippocampus | -0.167 [-0.224, -0.111] | 9.51e-09 * |
| Pallidum | -0.224 [-0.280, -0.167] | 1.72e-14 * |
| Putamen | -0.146 [-0.202, -0.089] | 5.52e-07 * |
| Caudate | -0.107 [-0.164, -0.051] | 2.31e-04 * |
| Thalamus | -0.364 [-0.421, -0.307] | 2.23e-35 * |
| Cerebellum Cortex | -0.078 [-0.134, -0.022] | 7.27e-03 |
| Cerebellum White Matter | -0.145 [-0.201, -0.088] | 6.87e-07 * |
| Inferior Lateral Ventricles | 0.307 [0.250, 0.364] | 9.33e-26 * |
| Lateral Ventricles | 0.334 [0.278, 0.391] | 3.40e-30 * |
| Cerebral Cortex | -0.120 [-0.177, -0.064] | 3.50e-05 * |
| Cerebral White Matter | -0.245 [-0.302, -0.188] | 4.77e-17 * |
| Cortical Thickness | -0.010 [-0.066, 0.047] | 7.36e-01 |
| Insula | -0.127 [-0.184, -0.071] | 1.25e-05 * |
| Transverse Temporal | 0.011 [-0.046, 0.067] | 7.12e-01 |
| Temporal Pole | -0.109 [-0.165, -0.052] | 1.84e-04 * |
| Frontal Pole | -0.044 [-0.100, 0.013] | 1.32e-01 |
| Supramarginal | -0.006 [-0.063, 0.050] | 8.26e-01 |
| Superior Temporal | 0.019 [-0.037, 0.075] | 5.16e-01 |
| Superior Parietal | 0.103 [0.046, 0.159] | 4.15e-04 * |
| Superior Frontal | -0.045 [-0.101, 0.012] | 1.24e-01 |
| Rostral Middle Frontal | -0.024 [-0.080, 0.032] | 4.08e-01 |
| Rostral Anterior Cingulate | -0.060 [-0.117, -0.004] | 3.85e-02 |
| Precuneous | -0.011 [-0.067, 0.045] | 7.08e-01 |
| Precentral | -0.036 [-0.093, 0.020] | 2.10e-01 |
| Posterior Cingulate | 0.057 [0.000, 0.113] | 5.18e-02 |
| Post Central | 0.112 [0.055, 0.168] | 1.26e-04 * |
| Pericalcarine | 0.067 [0.011, 0.124] | 2.09e-02 |
| Pars Triangularis | 0.015 [-0.041, 0.071] | 6.07e-01 |
| Pars Orbitalis | -0.010 [-0.066, 0.047] | 7.40e-01 |
| Pars Opercularis | -0.027 [-0.083, 0.030] | 3.58e-01 |
| Paracentral | 0.049 [-0.008, 0.105] | 9.47e-02 |
| Parahippocampal | -0.002 [-0.058, 0.055] | 9.55e-01 |
| Middle Temporal | -0.053 [-0.109, 0.004] | 7.06e-02 |
| Medial Orbitofrontal | -0.051 [-0.108, -0.005] | 7.87e-02 |
| Lingual | 0.009 [-0.048, 0.065] | 7.62e-01 |
| Lateral Orbitofrontal | -0.062 [-0.118, -0.005] | 3.37e-02 |
| Lateral Occipital | 0.044 [-0.012, 0.101] | 1.29e-01 |
| Isthmus Cingulate | -0.036 [-0.092, 0.021] | 2.19e-01 |
| Inferior Temporal | -0.037 [-0.094, 0.019] | 2.01e-01 |
| Inferior Parietal | 0.024 [-0.032, 0.081] | 4.02e-01 |
| Fusiform | -0.056 [-0.112, 0.000] | 5.38e-02 |
| Entorhinal | -0.089 [-0.146, -0.033] | 2.18e-03 |
| Cuneous | 0.071 [0.015, 0.128] | 1.43e-02 |
| Caudal Middle Frontal | -0.014 [-0.070, 0.042] | 6.29e-01 |
| Caudal Anterior Cingulate | -0.055 [-0.111, 0.001] | 5.86e-02 |
| BanksSts | -0.030 [-0.087, 0.026] | 2.97e-01 |

### Supplementary Table S24: Comparisons of all ROIs for Population B.

| Population B | Cohen's d [95% CI] | p-value |
|---|---|---|
| **Non-Psychiatric Controls (N=1382) vs. Mental Disorders (N=2764)** | | |
| CSF | 0.194 [0.133, 0.255] | 4.37e-09 * |
| Brain Stem | -0.210 [-0.271, -0.149] | 1.95e-10 * |
| 4th Ventricle | 0.153 [0.092, 0.214] | 3.48e-06 * |
| 3rd Ventricle | 0.381 [0.319, 0.442] | 2.11e-30 * |
| Ventral DC | -0.322 [-0.384, -0.261] | 2.29e-22 * |
| Nucleus Accumbens | -0.269 [-0.330, -0.208] | 4.52e-16 * |
| Amygdala | -0.283 [-0.344, -0.222] | 1.16e-17 * |
| Hippocampus | -0.181 [-0.242, -0.120] | 4.25e-08 * |
| Pallidum | -0.275 [-0.336, -0.214] | 9.71e-17 * |
| Putamen | -0.173 [-0.234, -0.112] | 1.65e-07 * |
| Caudate | -0.118 [-0.179, -0.057] | 3.29e-04 * |
| Thalamus | -0.394 [-0.456, -0.333] | 1.74e-32 * |
| Cerebellum Cortex | -0.117 [-0.178, -0.056] | 3.90e-04 * |
| Cerebellum White Matter | -0.156 [-0.217, -0.095] | 2.22e-06 * |
| Inferior Lateral Ventricles | 0.344 [0.283, 0.406] | 2.97e-25 * |
| Lateral Ventricles | 0.376 [0.314, 0.437] | 1.18e-29 * |
| Cerebral Cortex | -0.193 [-0.254, -0.132] | 5.08e-09 * |
| Cerebral White Matter | -0.315 [-0.376, -0.253] | 2.15e-21 * |
| Cortical Thickness | -0.044 [-0.105, 0.017] | 1.78e-01 |
| Insula | -0.129 [-0.190, -0.068] | 9.66e-05 * |
| Transverse Temporal | 0.007 [-0.054, 0.068] | 8.32e-01 |
| Temporal Pole | -0.145 [-0.206, -0.084] | 1.06e-05 * |
| Frontal Pole | -0.096 [-0.157, -0.035] | 3.62e-03 |
| Supramarginal | -0.043 [-0.104, 0.018] | 1.90e-01 |
| Superior Temporal | -0.031 [-0.092, 0.030] | 3.44e-01 |
| Superior Parietal | 0.072 [0.011, 0.133] | 2.81e-02 |
| Superior Frontal | -0.090 [-0.151, -0.029] | 6.30e-03 |
| Rostral Middle Frontal | -0.055 [-0.116, -0.006] | 9.37e-02 |
| Rostral Anterior Cingulate | -0.093 [-0.154, -0.032] | 4.90e-03 |
| Precuneous | -0.054 [-0.115, 0.007] | 1.00e-01 |
| Precentral | -0.067 [-0.128, -0.006] | 4.15e-02 |
| Posterior Cingulate | 0.049 [-0.012, 0.110] | 1.37e-01 |
| Post Central | 0.093 [0.032, 0.154] | 4.69e-03 |
| Pericalcarine | 0.071 [0.010, 0.132] | 3.16e-02 |
| Pars Triangularis | -0.010 [-0.071, 0.051] | 7.62e-01 |
| Pars Orbitalis | -0.007 [-0.068, 0.054] | 8.39e-01 |
| Pars Opercularis | -0.061 [-0.122, 0.000] | 6.53e-02 |
| Paracentral | 0.033 [-0.028, 0.094] | 3.12e-01 |
| Parahippocampal | 0.003 [-0.058, 0.064] | 9.22e-01 |
| Middle Temporal | -0.104 [-0.165, -0.043] | 1.59e-03 |
| Medial Orbitofrontal | -0.062 [-0.123, -0.001] | 5.88e-02 |
| Lingual | 0.006 [-0.055, 0.067] | 8.60e-01 |
| Lateral Orbitofrontal | -0.059 [-0.120, 0.002] | 7.20e-02 |
| Lateral Occipital | 0.031 [-0.030, 0.092] | 3.44e-01 |
| Isthmus Cingulate | -0.067 [-0.128, -0.006] | 4.15e-02 |
| Inferior Temporal | -0.064 [-0.124, -0.003] | 5.40e-02 |
| Inferior Parietal | -0.014 [-0.075, 0.047] | 6.65e-01 |
| Fusiform | -0.058 [-0.119, 0.003] | 7.81e-02 |
| Entorhinal | -0.120 [-0.181, -0.059] | 2.66e-04 * |
| Cuneous | 0.058 [-0.003, 0.119] | 7.76e-02 |
| Caudal Middle Frontal | -0.038 [-0.095, 0.027] | 2.98e-01 |
| Caudal Anterior Cingulate | -0.123 [-0.184, -0.062] | 2.02e-04 * |
| BanksSts | -0.086 [-0.147, -0.025] | 9.24e-03 |

### Supplementary Table S25: Comparisons of all ROIs for Population C.

| Population C | Cohen’s d [95% CI] | p-value |
|---|---|---|
| **Non-Psychiatric Controls (N=1376) vs. Mental Disorders (N=785)** | | |
| CSF | 0.264 [0.179, 0.349] | 4.09e-09 * |
| Brain Stem | -0.153 [-0.237, -0.068] | 6.56e-04 * |
| 4th Ventricle | 0.049 [-0.035, 0.133] | 2.74e-01 |
| 3rd Ventricle | 0.313 [0.228, 0.398] | 3.56e-12 * |
| Ventral DC | -0.230 [-0.314, -0.145] | 3.02e-07 * |
| Nucleus Accumbens | -0.180 [-0.265, -0.096] | 5.82e-05 * |
| Amygdala | -0.250 [-0.335, -0.165] | 2.54e-08 * |
| Hippocampus | -0.145 [-0.229, -0.060] | 1.25e-03 |
| Pallidum | -0.192 [-0.277, -0.108] | 1.77e-05 * |
| Putamen | -0.081 [-0.165, 0.003] | 7.03e-02 |
| Caudate | -0.092 [-0.176, -0.008] | 3.98e-02 |
| Thalamus | -0.298 [-0.383, -0.213] | 3.35e-11 * |
| Cerebellum Cortex | -0.168 [-0.252, -0.083] | 1.81e-04 * |
| Cerebellum White Matter | -0.149 [-0.233, -0.065] | 8.79e-04 * |
| Inferior Lateral Ventricles | 0.351 [0.266, 0.436] | 6.65e-15 * |
| Lateral Ventricles | 0.272 [0.187, 0.357] | 1.46e-09 * |
| Cerebral Cortex | -0.225 [-0.310, -0.140] | 5.28e-07 * |
| Cerebral White Matter | -0.246 [-0.331, -0.162] | 4.05e-08 * |
| Cortical Thickness | -0.180 [-0.264, -0.095] | 5.98e-05 * |
| Insula | -0.177 [-0.262, -0.093] | 7.67e-05 * |
| Transverse Temporal | -0.091 [-0.176, -0.007] | 4.15e-02 |
| Temporal Pole | -0.234 [-0.318, -0.149] | 1.90e-07 * |
| Frontal Pole | -0.176 [-0.260, -0.091] | 8.68e-05 * |
| Supramarginal | -0.139 [-0.224, -0.055] | 1.88e-03 |
| Superior Temporal | -0.151 [-0.235, -0.066] | 7.62e-04 * |
| Superior Parietal | -0.074 [-0.159, 0.010] | 9.61e-02 |
| Superior Frontal | -0.212 [-0.297, -0.127] | 2.28e-06 * |
| Rostral Middle Frontal | -0.175 [-0.260, -0.090] | 9.41e-05 * |
| Rostral Anterior Cingulate | -0.124 [-0.208, -0.039] | 5.65e-03 |
| Precuneous | -0.180 [-0.265, -0.096] | 5.83e-05 * |
| Precentral | -0.143 [-0.228, -0.059] | 1.37e-03 |
| Posterior Cingulate | -0.042 [-0.126, 0.042] | 3.48e-01 |
| Post Central | -0.049 [-0.133, 0.036] | 2.77e-01 |
| Pericalcarine | -0.005 [-0.089, 0.079] | 9.10e-01 |
| Pars Triangularis | -0.110 [-0.194, -0.025] | 1.41e-02 |
| Pars Orbitalis | -0.085 [-0.170, -0.001] | 5.66e-02 |
| Pars Opercularis | -0.158 [-0.242, -0.073] | 4.37e-04 * |
| Paracentral | -0.070 [-0.155, 0.014] | 1.17e-01 |
| Parahippocampal | -0.018 [-0.102, 0.066] | 6.89e-01 |
| Middle Temporal | -0.191 [-0.276, -0.107] | 1.97e-05 * |
| Medial Orbitofrontal | -0.132 [-0.216, -0.047] | 3.24e-03 |
| Lingual | -0.122 [-0.207, -0.038] | 6.31e-03 |
| Lateral Orbitofrontal | -0.121 [-0.206, -0.037] | 6.84e-03 |
| Lateral Occipital | -0.098 [-0.182, -0.013] | 2.90e-02 |
| Isthmus Cingulate | -0.159 [-0.243, -0.074] | 3.89e-04 * |
| Inferior Temporal | -0.134 [-0.218, -0.049] | 2.82e-03 |
| Inferior Parietal | -0.135 [-0.219, -0.050] | 2.67e-03 |
| Fusiform | -0.167 [-0.251, -0.082] | 1.98e-04 * |
| Entorhinal | -0.191 [-0.276, -0.107] | 1.97e-05 * |
| Cuneous | -0.063 [-0.147, 0.022] | 1.62e-01 |
| Caudal Middle Frontal | -0.139 [-0.223, -0.055] | 1.91e-03 |
| Caudal Anterior Cingulate | -0.145 [-0.230, -0.061] | 1.18e-03 |
| BanksSts | -0.147 [-0.232, -0.063] | 1.02e-03 |

**Supplementary Table S26: Comparison of findings between mental disorders subgroups and controls, with prior ENIGMA studies for Population A.** R=Right hemisphere, L=Left hemisphere, B=Bilateral.

| Population A | Cohen's d [95% CI] | p-value | ENIGMA studies: Cohen's d [95% CI] [Reference] |
|---|---|---|---|
| **Non-Psychiatric Controls (N=2005) vs. Substance Use Disorder (N=574)** | | | |
| Amygdala | -0.385 [-0.463, -0.307] | 6.58e-16 * | R: -0.041 [-0.110, 0.028] L: -0.055 [-0.124, 0.014] [11] |
| Hippocampus | -0.239 [-0.316, -0.161] | 4.95e-07 * | R: -0.081 [-0.150, -0.012] L: -0.087 [-0.156, -0.018] [11] |
| Insula | -0.173 [-0.250, -0.095] | 2.67e-04 * | R: -0.042 [-0.111, 0.027] L: -0.056 [-0.125, 0.013] [11] |
| Middle Temporal | -0.012 [-0.089, 0.065] | 7.99e-01 | R: -0.026 [-0.095, 0.043] L: -0.030 [-0.099, 0.034] [11] |
| Paracentral | -0.026 [-0.104, 0.051] | 5.77e-01 | R: -0.024 [-0.093, 0.045] L: -0.031 [-0.100, 0.038] [11] |
| Precentral | -0.074 [-0.152, 0.003] | 1.17e-01 | R: -0.042 [-0.111, 0.027] L: -0.039 [-0.108, 0.030] [11] |
| Supramarginal | 0.043 [-0.034, 0.121] | 3.58e-01 | R: -0.026 [-0.095, 0.043] L: -0.027 [-0.096, 0.042] [11] |
| **Non-Psychiatric Controls (N=1987) vs. Schizophrenia Spectrum Disorder (N=435)** | | | |
| Hippocampus | 0.281 [0.201, 0.361] | 1.18e-07 * | B: -0.46 [-0.58, -0.34] [12] |
| Amygdala | -0.080 [-0.160, 0.000] | 1.31e-01 | B: -0.31 [-0.43, -0.18] [12] |
| Thalamus | -0.072 [-0.152, 0.007] | 1.72e-01 | B: -0.31 [-0.44, -0.18] [12] |
| Nucleus Accumbens | -0.111 [-0.190, -0.031] | 3.65e-02 | B: -0.25 [-0.36, -0.14] [12] |
| Pallidum | -0.074 [-0.154, 0.006] | 1.62e-01 | B: 0.21 [0.04, 0.39] [12] |
| Lateral Ventricles | -0.187 [-0.267, -0.107] | 4.24e-04 * | B: 0.37 [0.25, 0.49] [12] |
| Cortical Thickness | -0.132 [-0.211, -0.052] | 1.30e-02 | R: -0.516 [-0.618, -0.414] L: -0.530 [-0.637, -0.423] [13] |
| Fusiform | -0.072 [-0.152, 0.007] | 1.71e-01 | R: -0.536 [-0.62, -0.452] L: -0.491 [-0.579, -0.403] [13] |
| Inferior Temporal | -0.230 [-0.310, -0.150] | 1.48e-05 * | R: -0.439 [-0.522, -0.355] L: -0.449 [-0.543, -0.356] [13] |
| Middle Temporal | -0.276 [-0.356, -0.196] | 2.04e-07 * | R: -0.379 [-0.472, -0.286] L: -0.444 [-0.549, -0.339] [13] |
| Superior Temporal | -0.391 [-0.471, -0.311] | 2.09e-13 * | R: -0.438 [-0.526, -0.351] L: -0.440 [-0.529, -0.352] [13] |
| Insula | -0.168 [-0.248, -0.089] | 1.40e-03 * | R: -0.406 [-0.497, -0.315] L: -0.408 [-0.494, -0.322] [13] |
| **Non-Psychiatric Controls (N=2047) vs. Depression (N=676)** | | | |
| Hippocampus | -0.074 [-0.149, 0.001] | 9.51e-02 | B: -0.144 [-0.225, -0.064] [14] |
| Amygdala | -0.170 [-0.245, -0.095] | 1.32e-04 * | B: -0.060 [-0.132, 0.011] [14] |
| Lateral Ventricles | 0.241 [0.165, 0.316] | 6.31e-08 * | B: 0.056 [-0.017, 0.129] [14] |
| Posterior Cingulate | 0.051 [-0.024, 0.126] | 2.51e-01 | R: -0.093 [-0.152, -0.034] L: -0.099 [-0.158, -0.040] [15] |

| | | | |
|---|---|---|---|
| Fusiform | -0.009 [-0.084, 0.066] | 8.39e-01 | R: -0.116 [-0.198, -0.033] L: -0.117 [-0.176, -0.058] [15] |
| Insula | -0.066 [-0.141, 0.009] | 1.39e-01 | R: -0.115 [-0.195, -0.035] L: -0.111 [-0.177, -0.045] [15] |
| Medial Orbitofrontal | -0.075 [-0.150, 0.000] | 9.23e-02 | R: -0.131 [-0.224, -0.039] L: -0.134 [-0.208, -0.059] [15] |
| Rostral Anterior Cingulate | -0.034 [-0.109, 0.041] | 4.40e-01 | R: -0.098 [-0.165, -0.031] L: -0.130 [-0.216, -0.044] [15] |
| **Non-Psychiatric Controls (N=2024) vs. Anxiety Disorder (N=524)** | | | |
| Putamen | -0.127 [-0.205, -0.049] | 9.76e-03 * | R: -0.158 [-0.235, -0.080] L: -0.141 [-0.219, -0.062] [16] |
| Pallidum | -0.138 [-0.216, -0.060] | 4.97e-03 * | R: 0.099 [0.021, 0.176] L: 0.129 [0.050, 0.208] [16] |
| Hippocampus | -0.046 [-0.124, 0.032] | 3.49e-01 | R: -0.060 [-0.137, 0.016] L: -0.084 [-0.161, -0.007] [16] |
| Amygdala | -0.129 [-0.206, -0.051] | 8.75e-03 * | R: -0.040 [-0.118, 0.037] L: -0.076 [-0.153, -0.001] [16] |

**Supplementary Table S27: Comparison of findings between mental disorders subgroups and controls, with prior ENIGMA studies for Population B.** R=Right hemisphere, L=Left hemisphere, B=Bilateral.

| Population B | Cohen's d [95% CI] | p-value | ENIGMA studies: Cohen's d [95% CI] [Reference] |
|---|---|---|---|
| **Non-Psychiatric Controls (N=1335) vs. Substance Use Disorder (N=569)** | | | |
| Amygdala | -0.405 [-0.496, -0.314] | 1.04e-15 * | R: -0.041 [-0.110, 0.028] L: -0.055 [-0.124, 0.014] [11] |
| Hippocampus | -0.278 [-0.369, -0.188] | 3.13e-08 * | R: -0.081 [-0.150, -0.012] L: -0.087 [-0.156, -0.018] [11] |
| Insula | -0.188 [-0.278, -0.098] | 1.78e-04 * | R: -0.042 [-0.111, 0.027] L: -0.056 [-0.125, 0.013] [11] |
| Middle Temporal | -0.079 [-0.169, 0.011] | 1.15e-01 | R: -0.026 [-0.095, 0.043] L: -0.030 [-0.099, 0.034] [11] |
| Paracentral | -0.055 [-0.145, 0.035] | 2.70e-01 | R: -0.024 [-0.093, 0.045] L: -0.031 [-0.100, 0.038] [11] |
| Precentral | -0.117 [-0.207, -0.027] | 1.96e-02 | R: -0.042 [-0.111, 0.027] L: -0.039 [-0.108, 0.030] [11] |
| Supramarginal | -0.012 [-0.102, 0.078] | 8.13e-01 | R: -0.026 [-0.095, 0.043] L: -0.027 [-0.096, 0.042] [11] |
| **Non-Psychiatric Controls (N=1326) vs. Schizophrenia Spectrum Disorder (N=435)** | | | |
| Hippocampus | 0.344 [0.250, 0.438] | 5.82e-10 * | B: -0.46 [-0.58, -0.34] [12] |
| Amygdala | -0.055 [-0.148, 0.039] | 3.22e-01 | B: -0.31 [-0.43, -0.18] [12] |
| Thalamus | -0.063 [-0.157, 0.030] | 2.51e-01 | B: -0.31 [-0.44, -0.18] [12] |
| Nucleus Accumbens | -0.144 [-0.237, -0.050] | 9.33e-03 | B: -0.25 [-0.36, -0.14] [12] |
| Pallidum | -0.109 [-0.202, -0.015] | 4.94e-02 | B: 0.21 [0.04, 0.39] [12] |
| Lateral Ventricles | -0.169 [-0.263, -0.075] | 2.27e-03 * | B: 0.37 [0.25, 0.49] [12] |
| Cortical Thickness | -0.184 [-0.277, -0.090] | 9.13e-04 * | R: -0.516 [-0.618, -0.414] L: -0.530 [-0.637, -0.423] [13] |
| Fusiform | -0.084 [-0.177, -0.010] | 1.29e-01 | R: -0.536 [-0.62, -0.452] L: -0.491 [-0.579, -0.403] [13] |
| Inferior Temporal | -0.217 [-0.311, -0.123] | 8.90e-05 * | R: -0.439 [-0.522, -0.355] L: -0.449 [-0.543, -0.356] [13] |
| Middle Temporal | -0.284 [-0.378, -0.190] | 3.11e-07 * | R: -0.379 [-0.472, -0.286] L: -0.444 [-0.549, -0.339] [13] |
| Superior Temporal | -0.402 [-0.496, -0.307] | 5.34e-13 * | R: -0.438 [-0.526, -0.351] L: -0.440 [-0.529, -0.352] [13] |
| Insula | -0.222 [-0.316, -0.128] | 6.15e-05 * | R: -0.406 [-0.497, -0.315] L: -0.408 [-0.494, -0.322] [13] |
| **Non-Psychiatric Controls (N=1372) vs. Depression (N=674)** | | | |
| Hippocampus | -0.094 [-0.181, -0.007] | 4.61e-02 | B: -0.144 [-0.225, -0.064] [14] |
| Amygdala | -0.197 [-0.284, -0.110] | 2.90e-05 * | B: -0.060 [-0.132, 0.011] [14] |
| Lateral Ventricles | 0.321 [0.233, 0.408] | 1.21e-11 * | B: 0.056 [-0.017, 0.129] [14] |
| Posterior Cingulate | 0.044 [-0.043, 0.131] | 3.50e-01 | R: -0.093 [-0.152, -0.034] L: -0.099 [-0.158, -0.040] [15] |
| Fusiform | -0.018 [-0.105, 0.069] | 7.05e-01 | R: -0.116 [-0.198, -0.033] L: -0.117 [-0.176, -0.058] [15] |
| Insula | -0.063 [-0.150, 0.024] | 1.81e-01 | R: -0.115 [-0.195, -0.035] L: -0.111 [-0.177, -0.045] [15] |

| | | | |
|---|---|---|---|
| Medial Orbitofrontal | -0.084 [-0.171, 0.003] | 7.37e-02 | R: -0.131 [-0.224, -0.039] L: -0.134 [-0.208, -0.059] [15] |
| Rostral Anterior Cingulate | -0.061 [-0.147, 0.026] | 1.97e-01 | R: -0.098 [-0.165, -0.031] L: -0.130 [-0.216, -0.044] [15] |
| **Non-Psychiatric Controls (N=1350) vs. Anxiety Disorder (N=524)** | | | |
| Putamen | -0.163 [-0.254, -0.072] | 1.55e-03 * | R: -0.158 [-0.235, -0.080] L: -0.141 [-0.219, -0.062] [16] |
| Pallidum | -0.212 [-0.302, -0.121] | 4.14e-05 * | R: 0.099 [0.021, 0.176] L: 0.129 [0.050, 0.208] [16] |
| Hippocampus | -0.066 [-0.157, 0.024] | 1.08e-01 | R: -0.060 [-0.137, 0.016] L: -0.084 [-0.161, -0.007] [16] |
| Amygdala | -0.164 [-0.255, -0.073] | 1.47e-03 * | R: -0.040 [-0.118, 0.037] L: -0.076 [-0.153, -0.001] [16] |

**Supplementary Table S28: Comparison of findings between mental disorders subgroups and controls, with prior ENIGMA studies for Population C.** R=Right hemisphere, L=Left hemisphere, B=Bilateral.

| Population C | Cohen's d [95% CI] | p-value | ENIGMA studies: Cohen's d [95% CI] [Reference] |
|---|---|---|---|
| **Non-Psychiatric Controls (N=1201) vs. Substance Use Disorder (N=119)** | | | |
| Amygdala | -0.391 [-0.500, -0.282] | 4.91e-05 * | R: -0.041 [-0.110, 0.028] L: -0.055 [-0.124, 0.014] [11] |
| Hippocampus | -0.365 [-0.474, -0.256] | 1.52e-04 * | R: -0.081 [-0.150, -0.012] L: -0.087 [-0.156, -0.018] [11] |
| Insula | -0.348 [-0.457, -0.239] | 3.05e-04 * | R: -0.042 [-0.111, 0.027] L: -0.056 [-0.125, 0.013] [11] |
| Middle Temporal | -0.275 [-0.384, -0.167] | 4.28e-03 * | R: -0.026 [-0.095, 0.043] L: -0.030 [-0.099, 0.034] [11] |
| Paracentral | -0.338 [-0.447, -0.229] | 4.53e-04 * | R: -0.024 [-0.093, 0.045] L: -0.031 [-0.100, 0.038] [11] |
| Precentral | -0.363 [-0.472, -0.254] | 1.65e-04 * | R: -0.042 [-0.111, 0.027] L: -0.039 [-0.108, 0.030] [11] |
| Supramarginal | -0.254 [-0.363, -0.146] | 8.25e-03 | R: -0.026 [-0.095, 0.043] L: -0.027 [-0.096, 0.042] [11] |
| **Non-Psychiatric Controls (N=1269) vs. Schizophrenia Spectrum Disorder (N=214)** | | | |
| Hippocampus | 0.232 [0.130, 0.334] | 1.74e-03 * | B: -0.46 [-0.58, -0.34] [12] |
| Amygdala | -0.073 [-0.175, 0.029] | 3.26e-01 | B: -0.31 [-0.43, -0.18] [12] |
| Thalamus | -0.095 [-0.197, 0.007] | 1.98e-01 | B: -0.31 [-0.44, -0.18] [12] |
| Nucleus Accumbens | -0.164 [-0.266, -0.062] | 2.69e-02 | B: -0.25 [-0.36, -0.14] [12] |
| Pallidum | -0.172 [-0.274, -0.070] | 2.00e-02 | B: 0.21 [0.04, 0.39] [12] |
| Lateral Ventricles | -0.147 [-0.249, -0.045] | 4.72e-02 | B: 0.37 [0.25, 0.49] [12] |
| Cortical Thickness | -0.128 [-0.230, -0.027] | 8.23e-02 | R: -0.516 [-0.618, -0.414] L: -0.530 [-0.637, -0.423] [13] |
| Fusiform | -0.162 [-0.264, -0.060] | 2.83e-02 | R: -0.536 [-0.62, -0.452] L: -0.491 [-0.579, -0.403] [13] |
| Inferior Temporal | -0.110 [-0.212, -0.009] | 1.35e-01 | R: -0.439 [-0.522, -0.355] L: -0.449 [-0.543, -0.356] [13] |
| Middle Temporal | -0.222 [-0.324, -0.119] | 2.77e-03 * | R: -0.379 [-0.472, -0.286] L: -0.444 [-0.549, -0.339] [13] |
| Superior Temporal | -0.256 [-0.359, -0.154] | 5.35e-04 * | R: -0.438 [-0.526, -0.351] L: -0.440 [-0.529, -0.352] [13] |
| Insula | -0.166 [-0.268, -0.064] | 2.46e-02 | R: -0.406 [-0.497, -0.315] L: -0.408 [-0.494, -0.322] [13] |
| **Non-Psychiatric Controls (N=1335) vs. Depression (N=179)** | | | |
| Hippocampus | 0.074 [-0.027, 0.175] | 3.51e-01 | B: -0.144 [-0.225, -0.064] [14] |
| Amygdala | -0.096 [-0.197, 0.005] | 2.28e-01 | B: -0.060 [-0.132, 0.011] [14] |
| Lateral Ventricles | 0.146 [0.045, 0.247] | 6.73e-02 | B: 0.056 [-0.017, 0.129] [14] |
| Posterior Cingulate | 0.024 [-0.077, 0.124] | 7.67e-01 | R: -0.093 [-0.152, -0.034] L: -0.099 [-0.158, -0.040] [15] |
| Fusiform | -0.005 [-0.106, 0.096] | 9.49e-01 | R: -0.116 [-0.198, -0.033] L: -0.117 [-0.176, -0.058] [15] |

| | | | |
|---|---|---|---|
| Insula | -0.087 [-0.188, 0.013] | 2.72e-01 | R: -0.115 [-0.195, -0.035] L: -0.111 [-0.177, -0.045] [15] |
| Medial Orbitofrontal | -0.085 [-0.186, 0.016] | 2.84e-01 | R: -0.131 [-0.224, -0.039] L: -0.134 [-0.208, -0.059] [15] |
| Rostral Anterior Cingulate | -0.067 [-0.168, 0.034] | 4.00e-01 | R: -0.098 [-0.165, -0.031] L: -0.130 [-0.216, -0.044] [15] |
| **Non-Psychiatric Controls (N=1289) vs. Anxiety Disorder (N=155)** | | | |
| Putamen | -0.155 [-0.258, -0.051] | 6.91e-02 | R: -0.158 [-0.235, -0.080] L: -0.141 [-0.219, -0.062] [16] |
| Pallidum | -0.195 [-0.298, -0.091] | 2.20e-02 | R: 0.099 [0.021, 0.176] L: 0.129 [0.050, 0.208] [16] |
| Hippocampus | -0.024 [-0.127, 0.079] | 7.78e-01 | R: -0.060 [-0.137, 0.016] L: -0.084 [-0.161, -0.007] [16] |
| Amygdala | -0.187 [-0.291, -0.084] | 2.79e-02 | R: -0.040 [-0.118, 0.037] L: -0.076 [-0.153, -0.001] [16] |
